# Assessing the Age Specificity of Infection Fatality Rates for COVID-19: Systematic Review, Meta-Analysis, and Public Policy Implications

**DOI:** 10.1101/2020.07.23.20160895

**Authors:** Andrew T. Levin, William P. Hanage, Nana Owusu-Boaitey, Kensington B. Cochran, Seamus P. Walsh, Gideon Meyerowitz-Katz

## Abstract

**Objective:** Determine age-specific infection fatality rates for COVID-19 to inform public health policies and communications that help protect vulnerable age groups.

**Methods:** Studies of COVID-19 prevalence were collected by conducting an online search of published articles, preprints, and government reports that were publicly disseminated prior to 18 September 2020. The systematic review encompassed 113 studies, of which 27 studies (covering 34 geographical locations) satisfied the inclusion criteria and were included in the meta-analysis. Age-specific IFRs were computed using the prevalence data in conjunction with reported fatalities four weeks after the midpoint date of the study, reflecting typical lags in fatalities and reporting. Meta-regression procedures in Stata were used to analyze the infection fatality rate (IFR) by age.

**Results:** Our analysis finds a exponential relationship between age and IFR for COVID-19. The estimated age-specific IFR is very low for children and younger adults (e.g., 0.002% at age 10 and 0.01% at age 25) but increases progressively to 0.4% at age 55, 1.4% at age 65, 4.6% at age 75, and 15% at age 85. Moreover, our results indicate that about 90% of the variation in population IFR across geographical locations reflects differences in the age composition of the population and the extent to which relatively vulnerable age groups were exposed to the virus.

**Discussion:** These results indicate that COVID-19 is hazardous not only for the elderly but also for middle-aged adults, for whom the infection fatality rate is two orders of magnitude greater than the annualized risk of a fatal automobile accident and far more dangerous than seasonal influenza. Moreover, the overall IFR for COVID-19 should not be viewed as a fixed parameter but as intrinsically linked to the age-specific pattern of infections. Consequently, public health measures to mitigate infections in older adults could substantially decrease total deaths.

## Introduction

Since the onset of the COVID-19 pandemic in winter 2020, it has been evident that the severity of the disease varies markedly across infected individuals.[1] Some remain asymptomatic throughout the course of infection or experience only mild symptoms such as headache or ageusia, whereas others experience much more severe illness, hospitalization, or even death.[2] Thus, official case reporting may tend to encompass a high fraction of severe cases but only a small fraction of asymptomatic or mildly symptomatic cases.[3] Moreover, the availability of live virus tests has varied significantly across locations and over time, and the deployment of such tests may differ markedly across demographic groups.

Consequently, assessments of the *case fatality rate (CFR)*, the ratio of deaths to reported cases, are fraught with pitfalls in gauging the severity of COVID-19. For example, early case reports from Wuhan noted a preponderance of older people among hospital admissions and a high CFR.[4] Subsequent studies have documented that children and young adults tend to exhibit fewer and milder symptoms and a far lower CFR than middle-aged and older adults.[5, 6] Nonetheless, the link between age and severity of COVID-19 infections has remained unclear for the reasons noted above.

To provide more accurate assessments of the spread of COVID-19, researchers have conducted seroprevalence studies in numerous locations.[7, 8] Such studies analyze samples of serum to detect antibodies in those infected with SARS-CoV-2, the virus that causes COVID-19. Estimates of prevalence (which includes asymptomatic and mildly symptomatic infections) can be used to estimate the *infection fatality rate (IFR)*, the ratio of fatalities to total infections, thereby facilitating the identification of vulnerable segments of the population and informing key policy decisions aimed at mitigating the consequences of the pandemic.[9]

For example, as shown in Table 1, the New York Department of Health conducted a large-scale seroprevalence study and estimated about 1·6 million SARS-CoV-2 infections among the 8 million residents of New York City.[10] However, only one-tenth of those infections were captured in reported COVID-19 cases, about one-fourth of which required hospitalization, and a substantial fraction of cases had fatal outcomes.[11] All told, COVID-19 fatalities in NYC represented a tenth of reported cases but only one-hundredth of all SARS-CoV-2 infections.

**Table 1:** COVID-19 in New York City during spring 2020.

|  | **<u>Number</u>** | **<u>Share of Infections</u>** |
| --- | --- | --- |
| NYC residents | 8 million | NA |
| Estimated infections | 1·6 million | 100% |
| Symptomatic infections | 1·1 million | 65% |
| Reported cases | 220 thousand | 12% |
| Hospitalized patients | 55 thousand | 3% |
| Fatal outcomes | 17 thousand | 1% |

*Note:* This table reports on the characteristics of COVID-19 infections in New York City (NYC). Infection prevalence was estimated by the New York Department of Health using samples collected on April 19-28, 2020.[10] The ratio of symptomatic to total infections reflects a recent assessment by the U.S. Center for Disease Control and Prevention.[38] The number of cases, hospitalized patients, and fatal outcomes is reported by the NYC Department of Health; as of May 22 (four weeks after the midpoint of the seroprevalence study), NYC had 17336 confirmed COVID-19 deaths.[11]

Nonetheless, divergences in study design and reporting have hampered comparisons of seroprevalence and IFRs across locations and demographic groups. For example, a number of studies have analyzed a representative sample of the general population, while other studies have made use of “convenience samples” of residual sera collected for other purposes (such as laboratory tests or blood donations).[12-14] Some studies have simply reported results for raw prevalence (the fraction of seropositive results), whereas other studies have reported results adjusted for antibody test characteristics (sensitivity and specificity).

While the NYC data indicate a population IFR of about 1%, seroprevalence estimates from other locations have yielded a wide array of population IFR estimates, ranging from about 0·6% in Geneva to levels exceeding 2% in northern Italy. Such estimates have fueled intense controversy about the severity of COVID-19 and the appropriate design of public health measures to contain it, which in turn hinges on whether the hazards of this disease are mostly limited to the elderly and infirm. Indeed, a recent meta-analysis noted the high degree of heterogeneity across aggregate estimates of IFR and concluded that research on age-stratified IFR is *“urgently needed to inform policymaking*.*”*[15]

This paper reports on a systematic review and meta-analysis of age-specific IFRs for COVID-19. We specifically consider the hypothesis that the observed variation in IFR across locations may primarily reflect the age specificity of COVID-19 infections and fatalities. Based on these findings, we are able to assess and contextualize the severity of COVID-19 and examine how age-specific prevalence affects the population IFR and the total incidence of fatalities.

## Methodology

To perform the present meta-analysis, we collected published papers and preprints on the seroprevalence and/or infection fatality rate of COVID-19 that were publicly disseminated prior to 18 September 2020. As described in Supplementary Appendix B, we systematically performed online searches in MedRxiv, Medline, PubMed, Google Scholar, and EMBASE, and we identified other studies listed in reports by government institutions such as the U.K. Parliament Office.[16] Data was extracted from studies by three authors and verified prior to inclusion.

We restricted our meta-analysis to studies of advanced economies, based on current membership in the Organization for Economic Cooperation and Development (OECD), in light of the distinct challenges of health care provision and reporting of fatalities in developing economies.[17] We also excluded studies aimed at measuring prevalence in specific groups such as health care workers.

Our meta-analysis encompasses two distinct approaches for assessing the prevalence of COVID-19: (1) seroprevalence studies that test for antibodies produced in response to the virus, and (2) comprehensive tracing programs using extensive live-virus testing of everyone who has had contact with a potentially infected individual. Seroprevalence estimates are associated with uncertainty related to the sensitivity and specificity of the test method and the extent to which the sampling frame provides an accurate representation of prevalence in the general population; see Supplementary Appendix C. Prevalence measures from comprehensive tracing programs are associated with uncertainty about the extent of inclusion of infected individuals, especially those who are asymptomatic.

### Sampling frame

To assess prevalence in the general population, a study should be specifically designed to utilize a random sample using standard survey procedures such as stratification and weighting by demographic characteristics. Other sampling frames may be useful for specific purposes such as sentinel surveillance but not well-suited for assessing prevalence due to substantial risk of systemic bias. Consequently, our meta-analysis excludes the following types of studies:

- *Blood Donors*. Only a small fraction of blood donors are ages 60 and above—a fundamental limitation in assessing COVID-19 prevalence and IFRs for older age groups—and the social behavior of blood donors may be systematically different from their peers.[13, 18] These concerns can be directly investigated by comparing alternative seroprevalence surveys of the same geographical location. As of early June, Public Health England (PHE) reported seroprevalence of 8·5% based on specimens from blood donors, whereas the U.K. Office of National Statistics (ONS) reported markedly lower seroprevalence of 5·4% (CI: 4·3–6·5%) based on its monitoring of a representative sample of the English population.[19, 20]
- *Dialysis Centers*. Assessing seroprevalence of dialysis patients using residual sera collected at dialysis centers is crucial for gauging the infection risks faced by these individuals, of which a disproportionately high fraction tend to be underrepresented minorities. Nonetheless, the seroprevalence within this group may be markedly different from that of the general population. For example, a study of U.K. dialysis patients found seroprevalence of about 36%, several times higher than that obtained using a very large random sample of the English population.[21, 22] Similarly, a recent U.S. study found a seropositive rate of 34% for dialysis patients in New York state that was more than twice as high as the seroprevalence in a random sample of New York residents.[10, 23]
- *Hospitals and Urgent Care Clinics*. Estimates of seroprevalence among current medical patients are subject to substantial bias, as evident from a pair of studies conducted in Tokyo, Japan: One study found 41 positive cases among 1071 urgent care clinic patients, whereas the other study found only two confirmed positive results in a random sample of nearly 2000 Tokyo residents (seroprevalence estimates of 3·8% vs. 0·1%).[24, 25]
- *Active Recruitment*. Soliciting participants is particularly problematic in contexts of low prevalence, because seroprevalence can be markedly affected by a few individuals who volunteer due to concerns about prior exposure. For example, a Luxembourg study obtained positive antibody results for 35 out of 1,807 participants, but nearly half of those individuals (15 of 35) had previously had a positive live virus test, were residing in a household with someone who had a confirmed positive test, or had direct contact with someone else who had been infected.[26]

Our critical review has also underscored the pitfalls of seroprevalence studies based on “convenience samples” of residual sera collected for other purposes. For example, two studies assessed seroprevalence of Utah residents during spring 2020. The first study analyzed residual sera from two commercial laboratories and obtained a prevalence estimate of 2·2% (CI: 1·2– 3·4%), whereas the second study collected specimens from a representative sample and obtained a markedly lower prevalence estimate of 0·96% (CI: 0·4–1·8%).[27, 28] In light of these issues, our meta-analysis includes residual serum studies but we flag such studies as having an elevated risk of bias.

### Comprehensive Tracing Programs

Our meta-analysis incorporates data on COVID-19 prevalence and fatalities in countries that have consistently maintained comprehensive tracing programs since the early stages of the pandemic. Such a program was only feasible in places where public health officials could conduct repeated tests of potentially infected individuals and trace those whom they had direct contact. We identify such countries using a threshold of 300 for the ratio of cumulative tests to reported cases as of 30 April 2020, based on comparisons of prevalence estimates and reported cases in Czech Republic, Korea, and Iceland; see Supplementary Appendices D and E.[29] Studies of Iceland and Korea found that estimated prevalence was moderately higher than the number of reported cases, especially for younger age groups; hence we make corresponding adjustments for other countries with comprehensive tracing programs, and we identify these estimates as subject to an elevated risk of bias.[30–32]

### Measurement of fatalities

Accurately measuring total deaths is a substantial issue in assessing IFR due to time lags from onset of symptoms to death and from death to official reporting. Symptoms typically develop within 6 days after exposure but may develop as early as 2 days or as late as 14 days.[1, 33] More than 95% of symptomatic COVID patients have positive antibody (IgG) titres within 17-19 days of symptom onset, and those antibodies remain elevated over a sustained period.[34–37] The mean time interval from symptom onset to death is 15 days for ages 18–64 and 12 days for ages 65+, with interquartile ranges of 9–24 days and 7–19 days, respectively, while the mean interval from date of death to the reporting of that person’s death is about 7 days with an IQR of 2–19 days; thus, the upper bound of the 95% confidence interval between symptom onset and reporting of fatalities is about six weeks (41 days).[38]

Figure 1 illustrates these findings in a hypothetical scenario where the pandemic was curtailed two weeks prior to the date of the seroprevalence study. This figure shows the results of a simulation calibrated to reflect the estimated distribution for time lags between symptom onset, death, and inclusion in official fatality reports. The histogram shows the frequency of deaths and reported fatalities associated with the infections that occurred on the last day prior to full containment. Consistent with the confidence intervals noted above, 95% of cumulative fatalities are reported within roughly four weeks of the date of the seroprevalence study.

**Figure 1:**
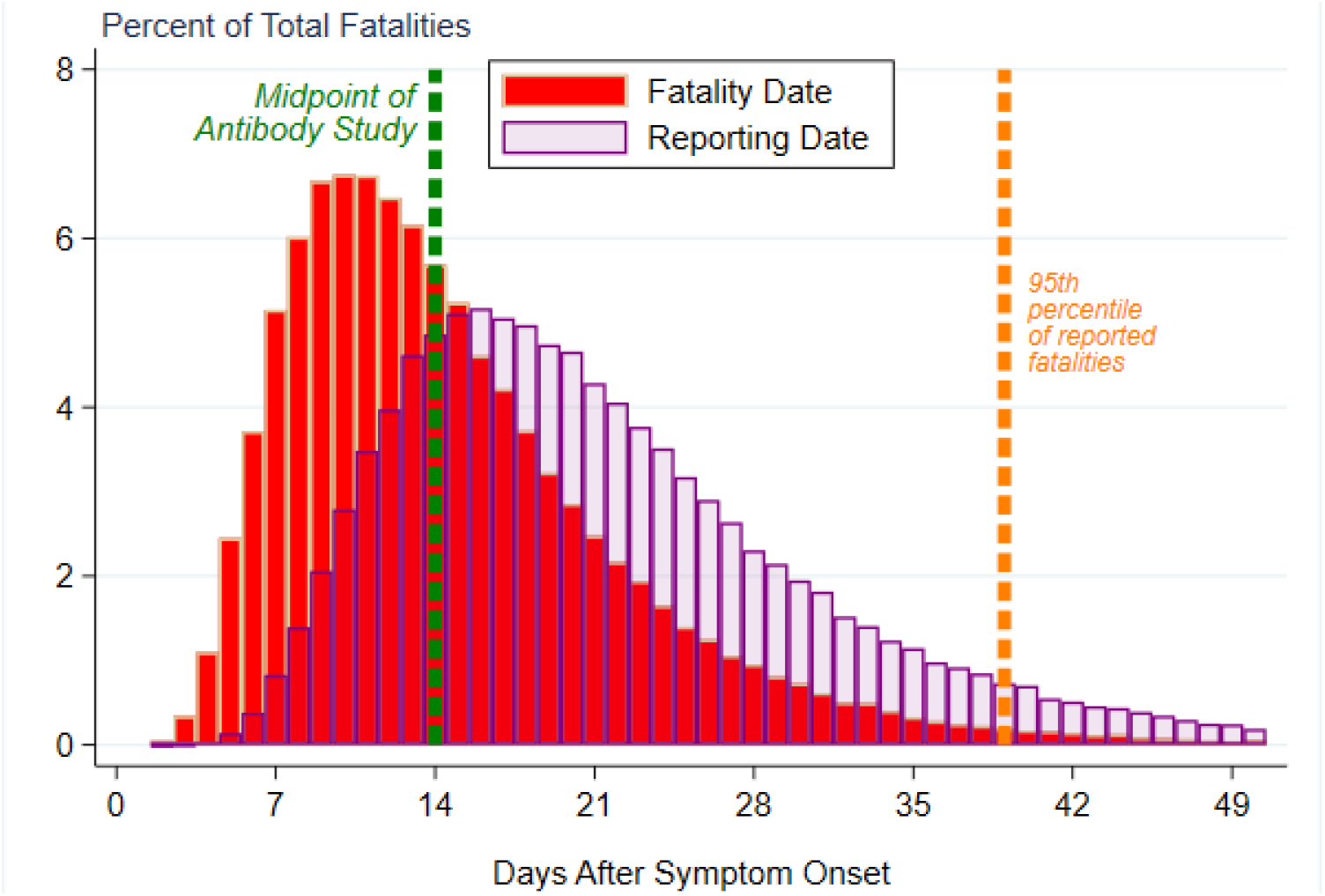
Time lags in the incidence and reporting of COVID-19 fatalities. *Note*: This figure illustrates time lags in the incidence and reporting of COVID-19 fatalities using the results of a simulation calibrated to reflect the estimated distribution for time lags between symptom onset, death, and inclusion in official fatality reports.[38] As indicated by the vertical green line, this simulation assumes that the seroprevalence study was conducted two weeks after the pandemic was curtailed. The histogram shows the frequency of deaths and reported fatalities associated with the infections that occurred on the last day prior to full containment. As indicated by the orange vertical line, 95% of cumulative fatalities are reported within about four weeks after the midpoint date of the seroprevalence study.

**Figure 2:**
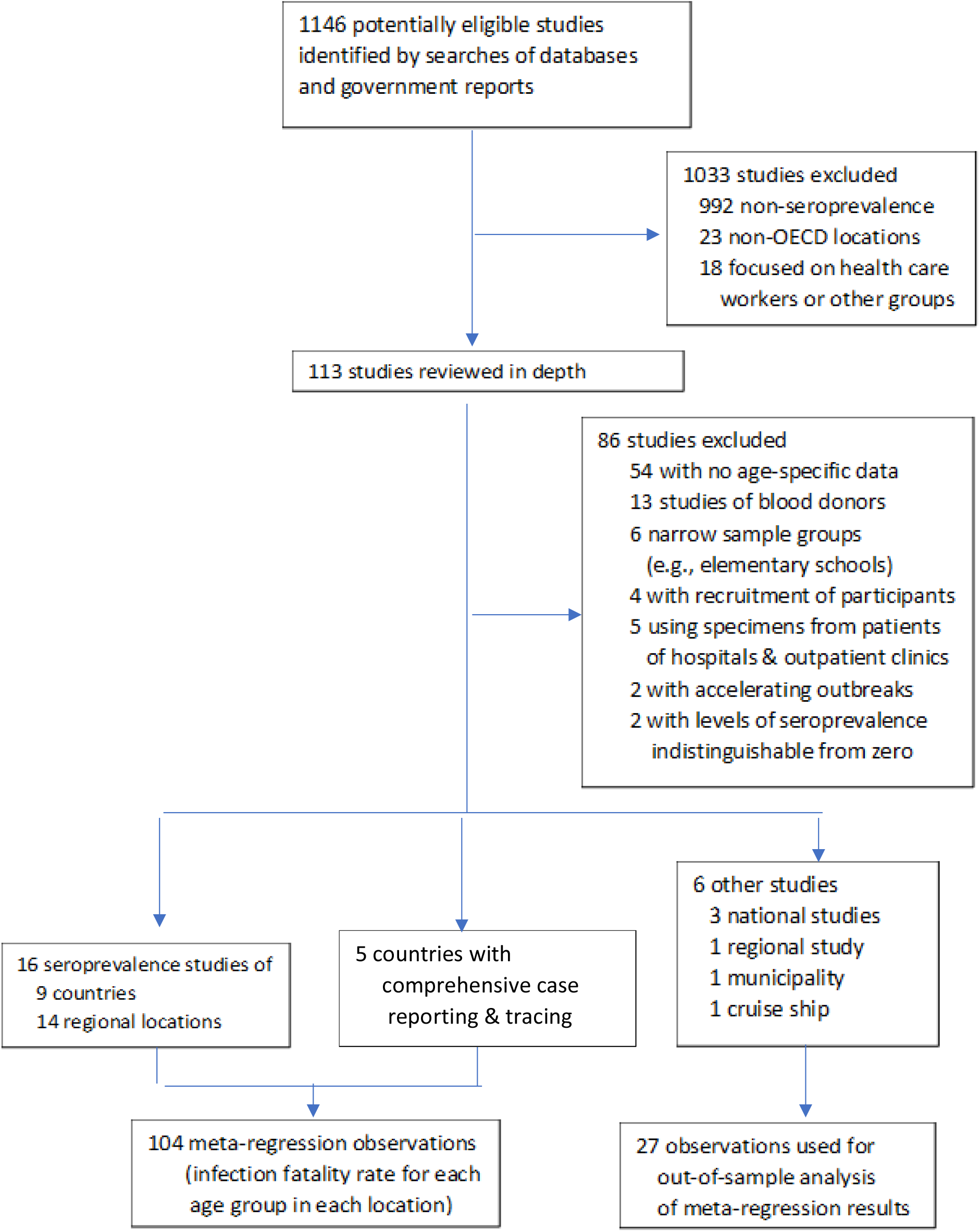
Study selection (PRISMA flow diagram)

As shown in Table 2, the precise timing of the count of cumulative fatalities is relatively innocuous in locations where the outbreak had been contained for more than a month prior to the date of the seroprevalence study. By contrast, in instances where the outbreak had only recently been contained, the death count continued rising markedly for several more weeks after the midpoint of the seroprevalence study.

**Table 2:** Timing of reported fatalities for selected seroprevalence studies.

| Location | Cumulative Fatalities |  |  | Change (%) |  |
| --- | --- | --- | --- | --- | --- |
|  | Study midpoint | 4 weeks later | 5 weeks later | Weeks 0 to 4 | Weeks 4 to 5 |
| Europe |  |  |  |  |  |
| Belgium | 6,262 | 8,843 | 9,150 | 41 | 3 |
| Geneva, Switzerland | 255 | 287 | 291 | 13 | 1 |
| Spain | 26,834 | 27,136 | 28,324 | 1 | 4 |
| Sweden | 2,586 | 3,831 | 3,940 | 48 | 3 |
| USA |  |  |  |  |  |
| Connecticut | 2,257 | 3,637 | 3,686 | 61 | 1 |
| Indiana | 932 | 1,984 | 2,142 | 113 | 8 |
| Louisiana | 477 | 2,012 | 2,286 | 322 | 14 |
| Miami | 513 | 1,160 | 1,290 | 126 | 11 |
| Minneapolis | 393 | 964 | 1093 | 145 | 13 |
| Missouri | 218 | 562 | 661 | 158 | 18 |
| New York | 20,212 | 28,663 | 29,438 | 42 | 3 |
| Philadelphia | 456 | 1509 | 1754 | 231 | 16 |
| San Francisco | 265 | 424 | 449 | 60 | 6 |
| Seattle | 536 | 732 | 775 | 37 | 6 |
| Utah | 41 | 96 | 98 | 134 | 2 |

*Note:* This table shows data on confirmed COVID-19 deaths for 4 European locations and 11 U.S. locations where seroprevalence has been assessed.[27, 124, 129, 163-165] Sources of fatality data are given in Supplementary Appendix F. For each location, the second column shows cumulative fatalities from the onset of the pandemic until the midpoint date of that seroprevalence study, while the next two columns report cumulative deaths 4 and 5 weeks later, respectively. The last two columns show the percent change in cumulative fatalities over each specified time interval (weeks 0 to 4 and weeks 4 to 5, respectively).

Therefore, we construct age-specific IFRs using the seroprevalence data in conjunction with cumulative fatalities four weeks after the midpoint date of each study; see Supplementary Appendix F. We have also conducted sensitivity analysis using cumulative fatalities five weeks after the midpoint date, and we flag studies as having an elevated risk of bias if the change in cumulative fatalities between weeks 4 and 5 exceeds 10%.

By contrast, matching prevalence estimates with subsequent fatalities is not feasible if a seroprevalence study was conducted in the midst of an accelerating outbreak. Therefore, our meta-analysis excludes seroprevalence studies for which the change in cumulative fatalities from week 0 to week 4 exceeds 200%.

### Metaregression procedure

To analyze IFR by age, we use meta-regression with random effects, using the *meta regress* procedure in *Stata* v16.[39, 40] In the metaregression, the dependent variable is the IFR for a specific age group in a specific geographical location, and the explanatory variable is the median age of that particular age group. We used a random-effects procedures to allow for residual heterogeneity between studies and across age groups by assuming that these divergences are drawn from a Gaussian distribution.

To assess the robustness of the metaregression results, we conduct several forms of sensitivity analysis:

- Analyze whether the metaregression coefficients exhibit any significant differences across three broad age categories (ages 0-34, 35-59, and 60+ years);
- Analyze whether the results are sensitive to exclusion of the oldest age group in each location (e.g., ages 65+ years), given that such groups may span a relatively wide age range and hence not fully capture their vulnerability to the virus;
- Conduct out-of-sample analysis using small-scale seroprevalence studies as well as studies not included in the metaregression due to overlapping geographical regions;
- Compare the actual population IFR in each location (based on estimated prevalence and confirmed deaths across all age groups) with the population IFR predicted by the metaregression (computed using the estimated prevalence and the predicted number of deaths within each reported age group).

Finally, publication bias is assessed using Egger’s regression and the trim-and-fill method.

## Results

After an initial screening of 1146 studies, we reviewed the full texts of 113 studies, of which 54 studies were excluded due to lack of age-specific data on COVID-19 prevalence or fatalities.[20, 24, 25, 41–91] Seroprevalence estimates for two locations were excluded because the outbreak was still accelerating during the period when the specimens were being collected and from two other locations for which age-specific seroprevalence was not distinguishable from zero.[27, 92–94] Studies of non-representative samples were excluded as follows: 13 studies of blood donors; 5 studies of patients of hospitals, outpatient clinics, and dialysis centers; 4 studies with active recruitment of participants, and 6 narrow sample groups such as elementary schools.[19, 23, 25, 26, 92, 95–117] Supplementary Appendix H lists all excluded studies.

Consequently, our meta-analysis encompasses 27 studies of 34 geographical locations, of which 28 are included in our metaregression and 6 are used for out-of-sample analysis. The metaregression observations can be categorized into three distinct groups:

- *Representative samples* from studies of England, France, Ireland, Italy, Netherlands, Portugal, Spain, Geneva (Switzerland), and four U.S. locations (Atlanta, Indiana, New York, and Salt Lake City).[10, 22, 28, 118–127]
- *Convenience samples* from studies of Belgium, Sweden, Ontario (Canada), and eight U.S. locations (Connecticut, Louisiana, Miami, Minneapolis, Missouri, Philadelphia, San Francisco, and Seattle).[27, 128–130]
- *Comprehensive tracing programs* for Australia, Iceland, Korea, Lithuania, and New Zealand.[131–135]

The metaregression includes results from the very large REACT-2 seroprevalence study of the English population.[22] Thus, to avoid pitfalls of nested or overlapping samples, two other somewhat smaller studies conducted by U.K. Biobank and the U.K. Office of National Statistics are not included in the metaregression but are instead used in out-of-sample analysis of the metaregression results.[20, 136] Similarly, the metaregression includes two large-scale studies involving representative samples from three French provinces and from Salt Lake City, and hence two other studies using convenience samples from laboratories in France and in Utah are used in the out-of-sample analysis along with two other small-scale studies.[27, 28, 137–139]

Data taken from included studies is shown in Supplementary Appendix I. Supplementary Appendix J assesses the risk of bias for each individual study. As indicated in Supplementary Appendix K, no publication bias was found using Egger’s test (*p* > 0.10), and the trim-and-fill method produced the same estimate as the metaregression.

We obtain the following metaregression results:

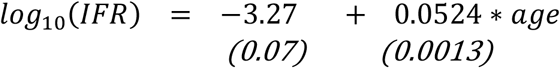

where the standard error for each estimated coefficient is given in parentheses. These estimates are highly significant with t-statistics of −44·5 and 40·4, respectively, and p-values below 0·0001. The residual heterogeneity τ^2^ = 0·071 (p-value < 0.0001) and I^2^ = 97·0, confirming that the random effects are essential for capturing unexplained variations across studies and age groups. The adjusted R^2^ is 94·7%.

As noted above, the validity of this metaregression rests on the condition that the data are consistent with a Gaussian distribution. The validity of that assumption is evident in Figure 3: Nearly all of the observations fall within the 95% prediction interval of the metaregression, and the remainder are moderate outliers.

**Figure 3:**
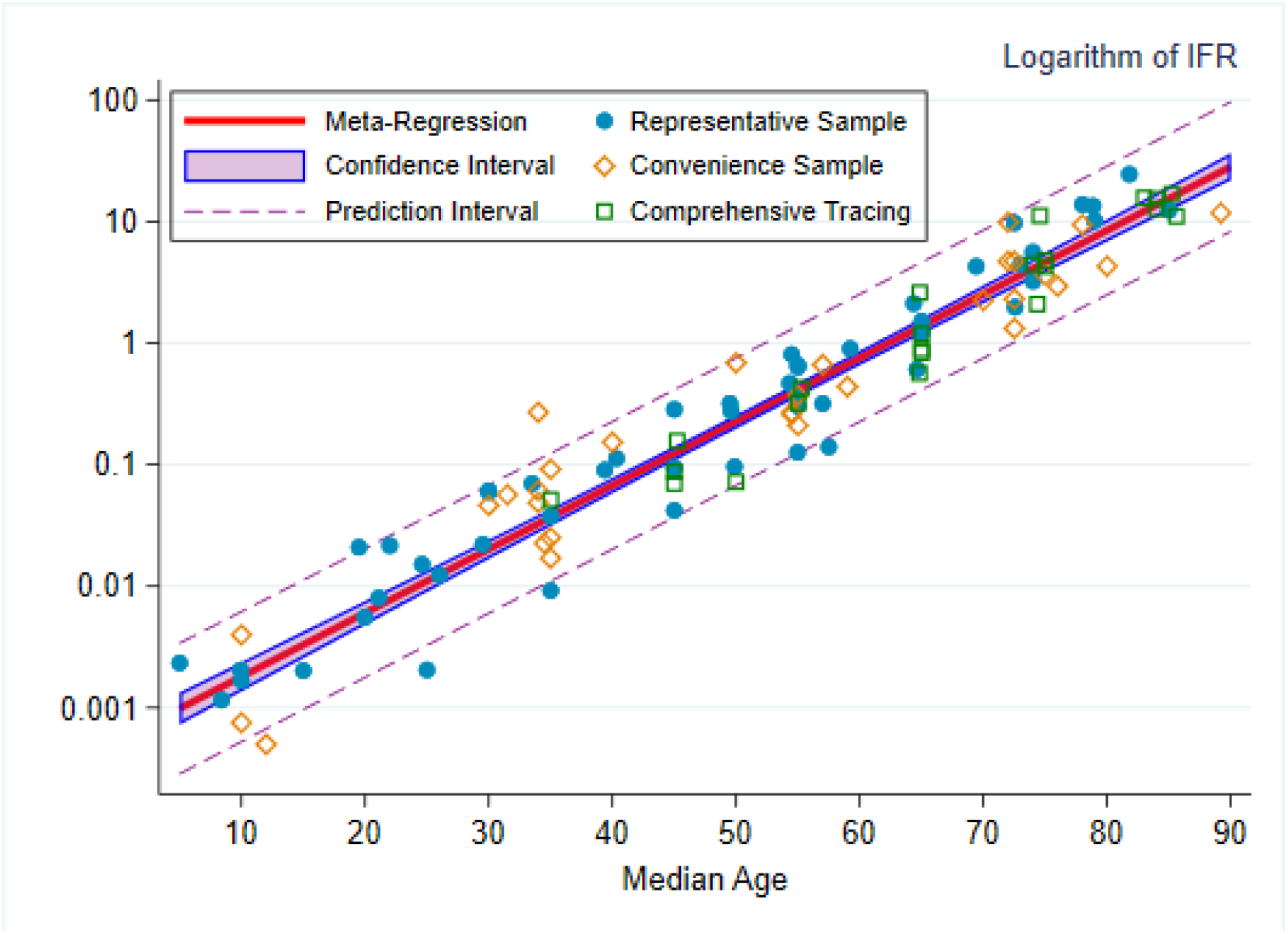
The log-linear relationship between IFR and age. *Note*: Our metaregression indicates that the infection fatality rate (IFR) increases exponentially with age, and hence this figure uses a base-10 logarithmic scale so that the relationship is evident across all ages from 5 to 95 years. Each marker denotes a specific metaregression observation, that is, the IFR for a particular age group in a particular location. The marker style reflects the type of observation: circles for observations from seroprevalence studies of representative samples, diamonds for seroprevalence studies of convenience samples, and squares for countries with comprehensive tracing programs. The red line denotes the metaregression estimate of IFR as a function of age, the shaded region depicts the 95% confidence interval for that estimate. The dashed lines denote the prediction interval (which includes random variations across studies and age groups), and almost all of the 108 metaregression observations lie within that interval.

This specification of the metaregression also assumes that the intercept and slope parameters are stable across the entire age distribution. We have confirmed the validity of that assumption by estimating alternative specifications in which the parameters are allowed to differ between three distinct age categories (ages 0–34, 35–59, and 60+ years). The estimated parameters are similar across all three age categories, and the null hypothesis of parameter constancy is consistent with the metaregression data. We have also confirmed that the metaregression results are not sensitive to exclusion of open-ended top age groups. (See Supplementary Appendix L for details.)

Figure 4 depicts the exponential relationship between age and the level of IFR in percent, and Figure 5 shows the corresponding forest plot. Evidently, the SARS-CoV-2 virus poses a substantial mortality risk for middle-aged adults and even higher risks for elderly people: The IFR is very low for children and young adults (e.g., 0.002% at age 10 and 0.01% at age 25) but rises to 0·4% at age 55, 1·4% at age 65, 4·6% at age 75, 15% at age 85, and exceeds 25% for ages 90 and above. These metaregression predictions are well aligned with the out-of-sample IFRs; see Supplementary Appendix M.

**Figure 4:**
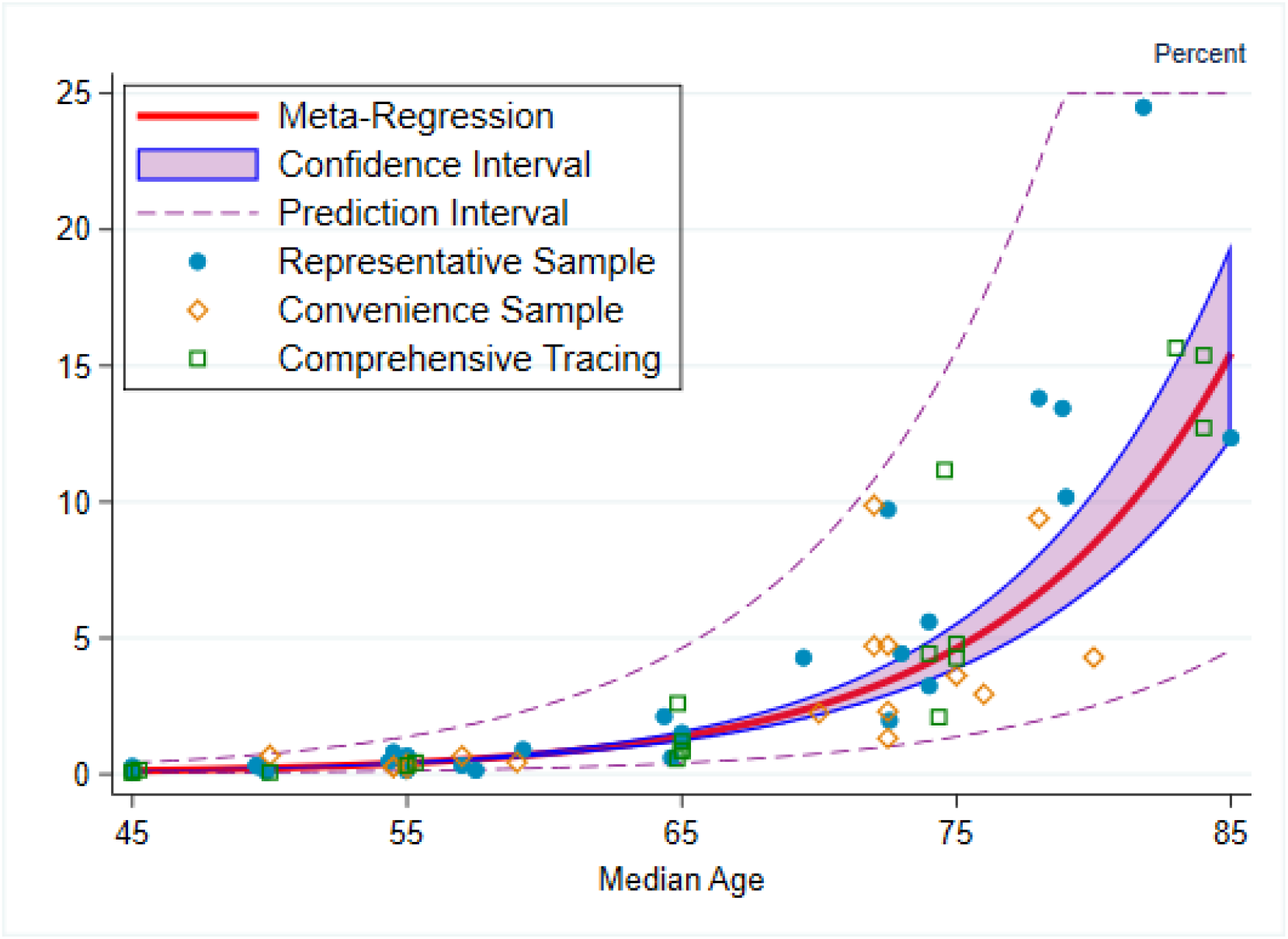
Benchmark analysis of the link between age and IFR. *Note*: This figure depicts the relationship between the infection fatality rate (IFR) and age, where IFR is shown in percentage terms. Each marker denotes a specific metaregression observation, that is, the IFR for a particular age group in a particular location. The marker style reflects the type of observation: circles for observations from seroprevalence studies of representative samples, diamonds for seroprevalence studies of convenience samples, and squares for countries with comprehensive tracing programs. The red line denotes the metaregression estimate of IFR as a function of age, the shaded region depicts the 95% confidence interval for that estimate. The dashed lines denote the prediction interval (which includes random variations across studies and age groups); almost all of the 104 metaregression observations lie within that interval.

**Figure 5:**
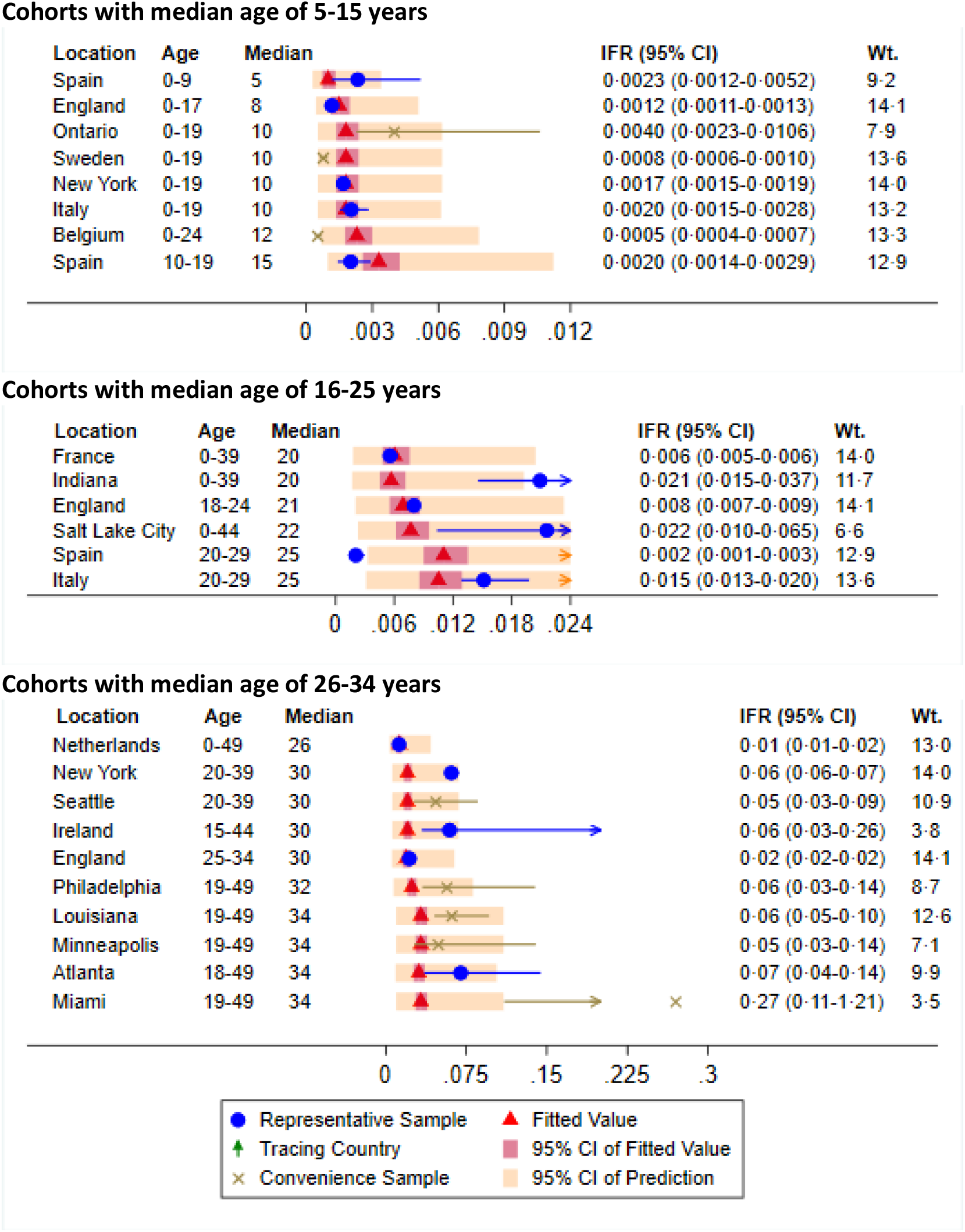
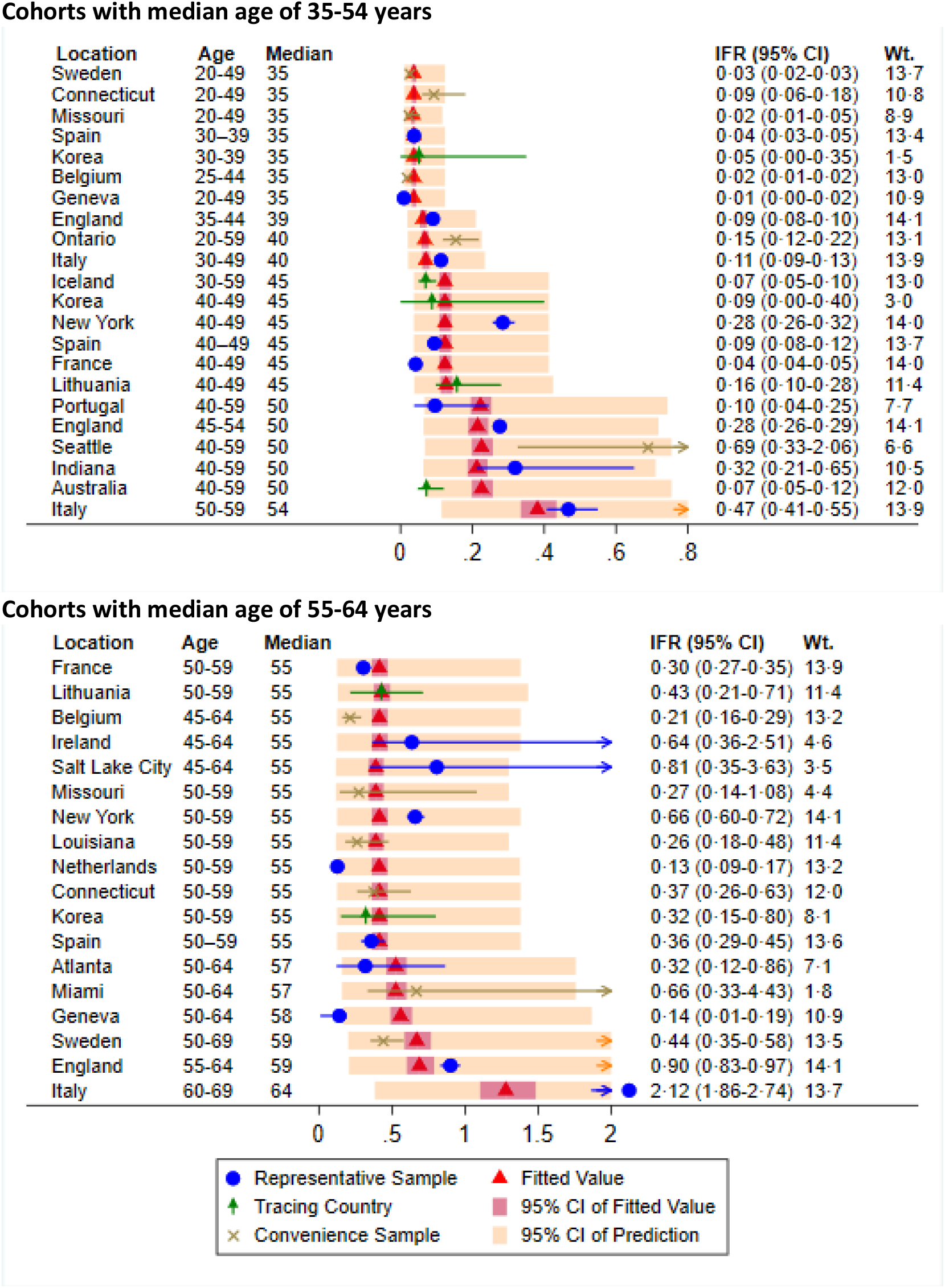
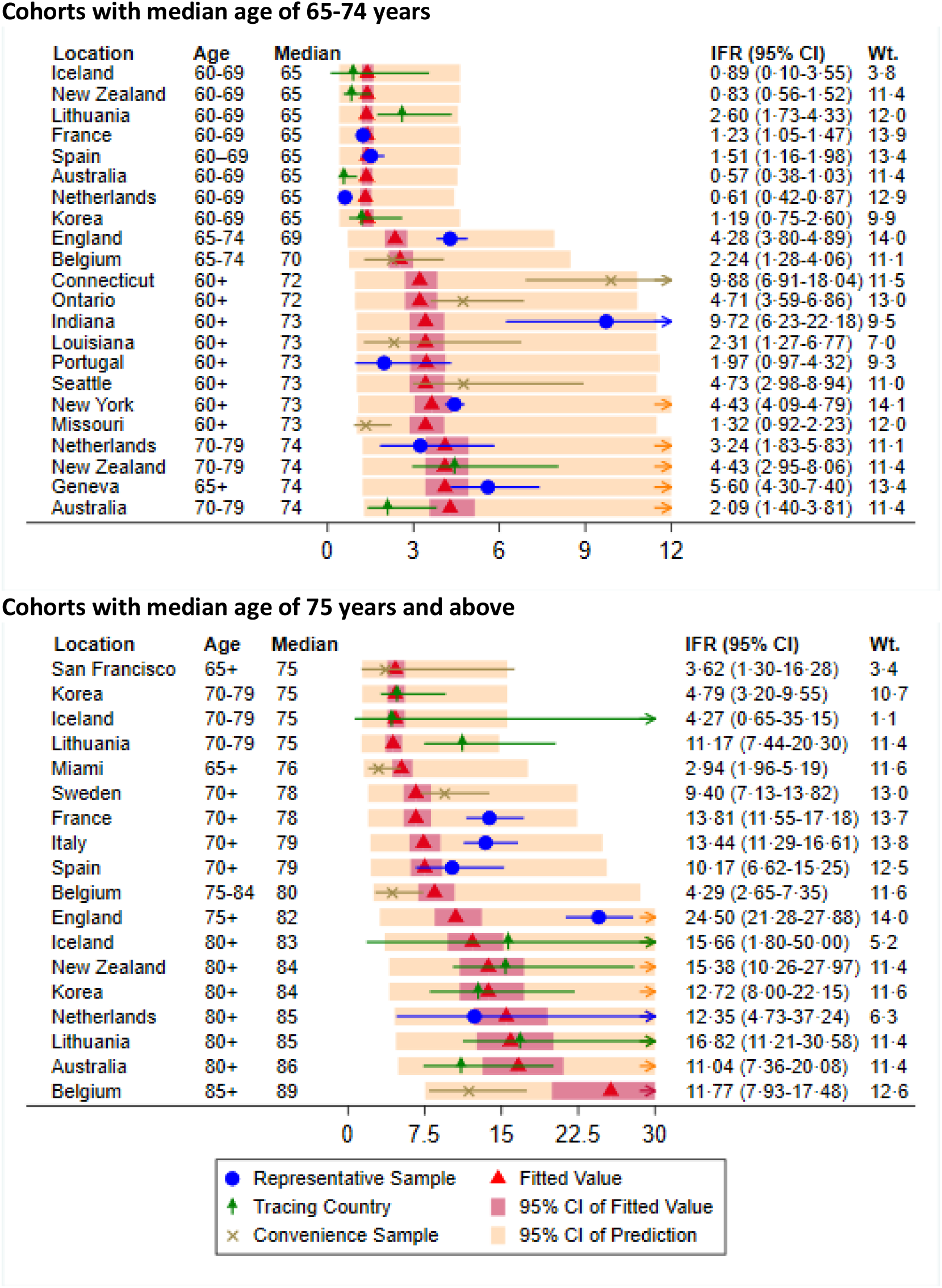
Forest plot of metaregression data.

As shown in Figure 6, population IFR (computed across all ages) ranges from about 0·5% in Salt Lake City and Geneva to 1·5% in Australia and England and 2·7% in Italy. The metaregression results indicate that about 90% of the variation in population IFR across geographical locations reflects differences in the age composition of the population and the extent to which relatively vulnerable age groups were exposed to the virus.

**Figure 6:**
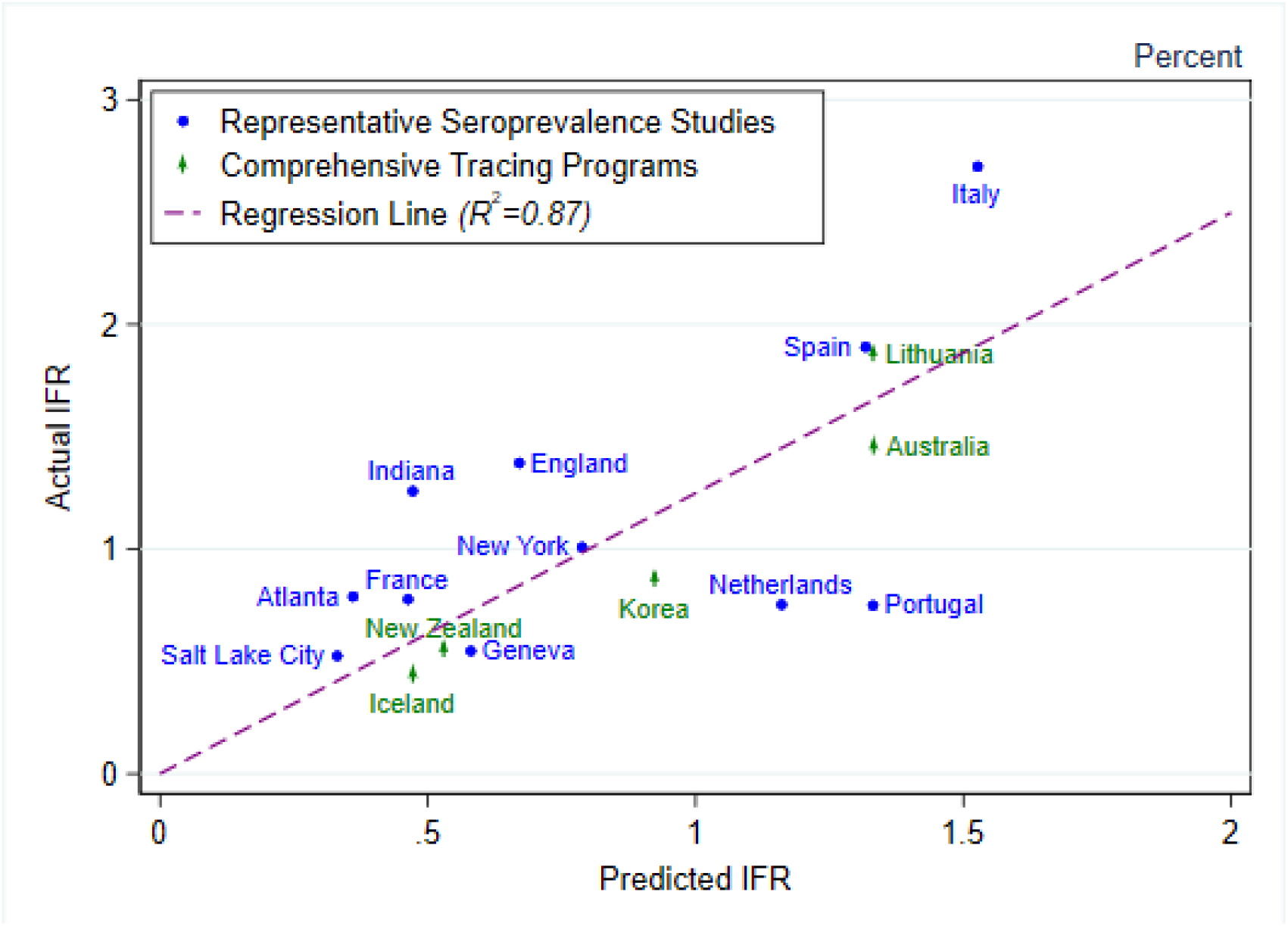
Variations in population IFR across geographical locations. *Note*: This figure depicts the extent to which the metaregression results account for variations in population IFR across geographical locations. The blue circles denote seroprevalence studies of representative samples, and the green diamonds denote countries with comprehensive tracing programs. For each observation, its position on the horizontal axis denotes its predicted IFR obtained by aggregating across the age-specific predictions of the metaregression, and its position on the vertical axis denotes the actual population IFR for that location. The dashed segments denote the estimated line obtained by fitting a regression to these 16 observations. The R^2^ of this regression is 0.87, indicating that nearly 90% of the variation in population IFR can be explained by variations in age composition and age-specific prevalence of COVID-19.

## Discussion

The IFR is central to our understanding of the public health impact of the COVID-19 pandemic and the appropriate policies for mitigating those consequences. In the absence of effective therapies or vaccines, such policies will primarily involve non-pharmaceutical interventions (NPIs). NPIs may include relatively mild measures (such as prohibitions on large gatherings) or more draconian restrictions such as shelter-in-place edicts, popularly known as “lockdowns.”

Unfortunately, public debate on these issues has been hampered by diverging assessments of the severity of COVID-19. For example, some early seroprevalence studies (using relatively small and non-representative samples, often in areas of low prevalence) yielded miniscule estimates of population IFR similar to those of seasonal influenza. Such estimates implied that strict NPIs would be completely irrational given the limited benefits and severe economic and social costs. With the dissemination of many more seroprevalence studies over recent months, a wide array of hypotheses have been mooted to explain the diverging implications for IFR, including regional variations in the quality of treatment or the extent of T-cell immunity to other betacoronaviruses.

By contrast, our critical review identifies the key characteristics of seroprevalence studies that can be used to provide reliable assessments of IFR. Indeed, once we focus on this group of studies (which includes nine national seroprevalence studies), our metaregression reveals a remarkably high degree of consistency in the implications for age-specific IFR. Moreover, our results indicate that most of the variation in population IFR across locations reflects differences in the extent to which vulnerable age groups were exposed to the virus.

One key implication of our findings is that the incidence of fatalities from a COVID-19 outbreak depends crucially on the age groups that are infected, which in turn reflects the age structure of that population and the extent to which public health measures limit the incidence of infections among vulnerable age groups.[140] Indeed, even if an outbreak is mainly concentrated among younger people, it may be very difficult to prevent the virus from spreading among older adults.[141]

To gauge the benefits of age-stratified public health strategies for COVID-19, we have constructed two illustrative scenarios for the U.S. trajectory of infections and fatalities (see Supplementary Appendix N). Each scenario assumes that U.S. prevalence rises to a plateau of around 20% but with different patterns of age-specific prevalence. In particular, if prevalence becomes uniform across age groups, this analysis projects that total U.S. fatalities would rise to nearly 900 thousand and that population IFR would converge to around 1·3%. By contrast, a scenario with relatively low incidence of new infections among vulnerable age groups would be associated with a much lower number of fatalities (about 350 thousand) and a correspondingly lower population IFR of about 0·5%.

A further implication of our results is that the risks of infection to the middle aged cannot be neglected. This is important for pandemic management strategies that aim to avoid large influxes of patients to healthcare. Indeed, it is likely that an unmitigated outbreak among middle-aged and older adults could have severe consequences on the healthcare system.

Public health communications can be helpful for persuading individuals to take steps to mitigate the risk of infection for themselves as well as others with whom they have direct contact (family members, friends, and colleagues). For this purpose, it is helpful to contextualize the magnitude of age-specific IFRs for COVID-19 relative to annualized fatality rates for other routine activities; that annual timeframe reflects the premise that effective vaccines and/or treatments for COVID-19 would hopefully become widely available sometime within the next year or two.

In particular, Table 3 compares the age-specific IFRs from our meta-regression analysis to the annualized risks of fatal automobile accidents or other unintentional injuries in England and in the United States.[142, 143] For example, an English person aged 55–64 years who gets infected with SARS-CoV-2 faces a fatality risk that is more than 200 times higher than the annual risk of dying in a fatal car accident. These results also confirm that COVID-19 is far more deadly than seasonal flu; indeed, the World Health Organization indicates that seasonal influenza mortality is usually well below 0·1% unless access to health care is constrained.[144] (See Supplementary Appendix O for further details.) Moreover, mortality from seasonal influenza outbreaks is mitigated by prior immunity and vaccinations, whereas that is not the case for SARS-CoV-2.

**Table 3:** Age-specific fatality rates for COVID-19 infections vs. accidental deaths (%)

| Age Group | COVID-19 IFR<br>(95% CI) | <u>Automobile Fatalities</u> |  | <u>Other Accidental Fatalities</u> |  |
| --- | --- | --- | --- | --- | --- |
|  |  | England | USA | England | USA |
| 0 to 34 | 0.004<br>(0.003–0.005) | 0.002 | 0.015 | 0.004 | 0.032 |
| 35 to 44 | 0.068<br>(0.058–0.078) | 0.002 | 0.012 | 0.017 | 0.043 |
| 45 to 54 | 0.23<br>(0.20–0.26) | 0.002 | 0.013 | 0.019 | 0.043 |
| 55 to 64 | 0.75<br>(0.66–0.87) | 0.003 | 0.013 | 0.014 | 0.043 |
| 65 to 74 | 2.5<br>(2.1–3.0) | 0.003 | 0.013 | 0.020 | 0.040 |
| 75 to 84 | 8.5<br>(6.9–10.4) | 0.005 | 0.017 | 0.069 | 0.094 |
| 85+ | 28.3<br>(21.8–36.6) | 0.007 | 0.019 | 0.329 | 0.349 |

*Note:* This table compares IFRs for COVID-19 with the incidence of accidental deaths in England and in the USA. For each age group, the second column shows the metaregression estimate of the age-specific IFR with its 95% confidence interval enclosed in parentheses. The final four columns report the annual incidence of automobile fatalities and other accidental fatalities as a percent of the population of each age group in each country. The accidental fatality data for England as of 2019 is reported by the U.K. Office of National Statistics, while the corresponding U.S. data are reported as of 2018 by the U.S. National Center for Health Statistics.[142, 143]

Our critical review highlights the benefits of assessing prevalence using large-scale studies of representative samples of the general population rather than convenience samples of blood donors or medical patients. Conducting such studies on an ongoing basis will enable public health officials to monitor changes in prevalence among vulnerable age groups and gauge the efficacy of public policy measures. Moreover, such studies enable researchers to assess the extent to which antibodies to SARS-CoV-2 may gradually diminish over time as well as the extent to which advances in treatment facilitate the reduction of age-specific IFRs.

Our critical review also underscores the importance of methodological issues in assessing IFR. For example, the raw prevalence results reported by a national study of Italy would imply a population IFR of about 2·3%, whereas test-adjusted prevalence implies a substantially higher IFR of 2·7%. Likewise, a few recent studies have excluded all deaths occurring in nursing homes and retirement communities and have obtained estimates of population IFR that are markedly lower than our estimates based on all confirmed COVID-19 fatalities, whereas assessments of IFR based on measures of excess mortality are broadly similar to our estimates.[120, 145–147] See Supplementary Appendix P for further discussion.

Our metaregression results are generally consistent with the study of Verity et al. (2020) and Ferguson et al. (2020), which were completed at an early stage of the COVID-19 pandemic and characterized an exponential pattern of age-specific IFRs (see Supplementary Appendix Q).[148, 149] Other subsequent studies have obtained broadly similar patterns of age-specific IFRs using statistical models to describe the dynamics of transmission and mortality using surveillance data in specific locations.[69, 150]

Our findings are well-aligned with a recent meta-analysis of population IFR and indeed explain a high proportion of the dispersion in population IFRs highlighted by that study.[151] In contrast, our findings are markedly different from those of another review of population IFR that includes samples that did not satisfy our inclusion criteria[152].

The exponential pattern of our age-specific IFR estimates is qualitatively similar to that of case fatality rates (CFRs). However, the relative magnitudes are systematically different, reflecting the extent to which asymptomatic or mildly symptomatic cases are much more common in younger adults than in middle-aged and older adults. For example, the ratio of CFR to IFR is about 15:1 for ages 30–49, about 7:1 for ages 50–69, and about 5:1 for ages 70–79 years (see Supplementary Appendix R).

A potential concern about measuring IFR based on seroprevalence is that antibody titers may diminish over time, leading to underestimation of true prevalence and corresponding overestimation of IFR. This concern is particularly relevant for seroprevalence studies conducted many months after the outbreak was contained.[153] However, recent research has confirmed that production of spike-specific antibodies to SARS-CoV-2 persists in a very high proportion of individuals for at least 3 or 4 months, which is the relevant timeframe for nearly all of the seroprevalence studies used in our metaregression.[91, 154] Moreover, one recent study has demonstrated that incorporating seroreversion into measures of prevalence does not have a material impact on estimates of age-specific IFRs.[155]

Moreover, a key feature of our metaregression analysis is that we also utilize age-specific IFR data based on RT-PCR results (*not* seroprevalence) for five countries that have maintained comprehensive tracing programs since the onset of the pandemic, namely, Australia, Iceland, Korea, Lithuania, and New Zealand. As shown in figure 3, the age-specific IFRs for those five countries are well aligned with the metaregression predictions, indicating that these findings do not rely upon any specific method of gauging prevalence.

A substantial limitation of our work is that we have not considered factors apart from age that affect the IFR of COVID-19. For example, we have not considered the extent to which IFRs may vary with demographic factors such as race and ethnicity or potential causal interactions between these factors.[41, 70] Likewise, our metaregression does not include measures of comorbidities such as diabetes or obesity.[156] However, a recent study of data from a large representative and longitudinal sample collected by U.K. Biobank found that measures of frailty and comorbidity had only moderate effects in predicting COVID-19 mortality risk (i.e., increased odds of about 10%); moreover, that link was negligible among positive COVID-19 cases after accounting for age and sex.[157] See Supplementary Appendix S for additional evidence. Further research on these issues is clearly warranted.

Another limitation of our meta-analysis is that we have focused exclusively on assessing IFRs in advanced economies to facilitate comparability regarding health care provision and reporting of fatalities. Nonetheless, it is absolutely clear that the COVID-19 pandemic has had devastating consequences for lower-income and developing countries. For example, as of late October 2020, the reported COVID-19 death counts were nearly 160 thousand in Brazil, 120 thousand in India, and 90 thousand in Mexico. And in many countries, measures of excess mortality are much higher than official tabulations of COVID-19 fatalities.

Consequently, analysis of prevalence and IFR is urgently needed to provide guidance to public health officials in developing countries.[155, 158, 159] However, the core findings of our meta-analysis may well be relevant even in those contexts. For example, recent prevalence studies of Manaus, Brazil found that about 66% of the population was infected with the SARS-CoV-2 virus between March and August 2020.[160, 161] As of October, Manaus (a city with 1·8 million inhabitants) had 2853 confirmed COVID-19 deaths.[162] That outcome is remarkably consistent with our analysis, because nearly 90% of Manaus residents are under 50 years of age. Indeed, using the age structure of the Manaus population and assuming uniform prevalence of infections across age groups, our metaregression predicts a population IFR of 0·22% that is practically indistinguishable from the observed outcome of 0·2%. (See Supplementary Appendix T.) Thus, our analysis provides a coherent explanation why Manaus was much less severely impacted by the pandemic compared to other locations with larger numbers of middle-aged and older adults.

Finally, it should be noted that our analysis has focused on assessing fatality risks but has not captured the full spectrum of adverse health consequences of COVID-19, some of which may be very severe and persistent. Further research is needed to assess age-stratified rates of hospitalization as well as longer-term sequelae attributable to SARS-CoV-2 infections. These factors are likely to be particularly important in quantifying overall risks to health care.

In summary, our analysis demonstrates that COVID-19 is not only dangerous for the elderly and infirm but also for healthy middle-aged adults. The metaregression explains nearly 90% of the geographical variation in population IFR, indicating that the population IFR is intrinsically linked to the age-specific pattern of infections. Consequently, public health measures to protect vulnerable age groups could substantially reduce the incidence of mortality.

## Data Availability

This study is a meta-analysis using information from published articles, preprints, and government reports; all sources are listed in the bibliography with active URLs. The data and Stata code used in performing the meta-regression analysis are provided as Supplementary Materials.

