## Supplementary Appendices for "Assessing the Age Specificity of Infection Fatality Rates for COVID-19: Systematic Review, Meta-Analysis, and Public Policy Implications"

#### Supplementary Appendix

Andrew T. Levin, William P. Hanage, Nana Owusu-Boaitey,
Kensington B. Cochran, Seamus P. Walsh, and Gideon Meyerowitz-Katz

30 October 2020

**Corresponding Author:**

Gideon Meyerowitz-Katz
Research, Monitoring & Surveillance Coordinator
Western Sydney Local Health District
PO Box 792
Seven Hills, NSW 2147 Australia


### List of Supplementary Appendices

**A: PRISMA Checklist**

**B: Meta-analysis search procedure**

**C: Seroprevalence adjustments for test characteristics**

**D: Countries with comprehensive tracing programs**

**E: Prevalence vs reported cases in Iceland**

**F: Age-specific fatality data and source information**

**G: Metaregression methodology**

**H: List of excluded studies**

**I: Seroprevalence Rates for Studies Included in Meta-Analysis**

**J: Assessment of risk of bias for included studies**

**K: Assessment of publication bias**

**L: Stability of metaregression results across age categories**

**M: Out-of-sample analysis of metaregression results**

**N: U.S. scenario analysis**

**O: Infection fatality rate for seasonal influenza**

**P: Comparison of confirmed fatalities and excess mortality**

**Q: Comparison of age-specific IFRs**

**R: Comparison of age-specific CFR vs. IFR**

**S: Comorbidities and demographic factors**

**T: Metaregression Predictions for Manaus, Brazil**

**Supplementary Appendix A: PRISMA Checklist**[1]

| Section/topic | # | Checklist item | Page # |
|---|---|---|---|
| **TITLE** | | | |
| Title | 1 | Identify the report as a systematic review, meta-analysis, or both. | Title Page |
| **ABSTRACT** | | | |
| Structured summary | 2 | Provide a structured summary including, as applicable: background; objectives; data sources; study eligibility criteria, participants, and interventions; study appraisal and synthesis methods; results; limitations; conclusions and implications of key findings; systematic review registration number. | Title Page |
| **INTRODUCTION** | | | |
| Rationale | 3 | Describe the rationale for the review in the context of what is already known. | 1-2 |
| Objectives | 4 | Provide an explicit statement of questions being addressed with reference to participants, interventions, comparisons, outcomes, and study design (PICOS). | 1-2 |
| **METHODS** | | | |
| Protocol and registration | 5 | Indicate if a review protocol exists, if and where it can be accessed (e.g., Web address), and, if available, provide registration information including registration number. | NA |
| Eligibility criteria | 6 | Specify study characteristics (e.g., PICOS, length of follow-up) and report characteristics (e.g., years considered, language, publication status) used as criteria for eligibility, giving rationale. | 2 |
| Information sources | 7 | Describe all information sources (e.g., databases with dates of coverage, contact with study authors to identify additional studies) in the search and date last searched. | 2, App.B |
| Search | 8 | Present full electronic search strategy for at least one database, including any limits used, such that it could be repeated. | App. B |
| Study selection | 9 | State the process for selecting studies (i.e., screening, eligibility, included in systematic review, included in the meta-analysis). | 2-5, App. D |
| Data collection process | 10 | Describe method of data extraction from reports (e.g., piloted forms, independently, in duplicate) and any processes for obtaining and confirming data from investigators. | 2, App. F, H |
| Data items | 11 | List and define all variables for which data were sought (e.g., PICOS, funding sources) and any assumptions and simplifications made. | 2-5 |
| Risk of bias in individual studies | 12 | Describe methods used for assessing risk of bias of individual studies and how this information is to be used in any data synthesis. | 2-5 |
| Summary measures | 13 | State principal summary measures. | 1-5 |
| Synthesis of results | 14 | Describe the methods of handling data and combining results of studies, if done, including measures of consistency. | 5, App. G |
| Risk of bias across studies | 15 | Specify any assessment of risk of bias that may affect the cumulative evidence (e.g., publication bias, selective reporting within studies). | 5, App. G |
| Additional analyses | 16 | Describe methods of additional analyses (e.g., sensitivity or subgroup analyses, meta-regression), if done, indicating which were pre-specified. | 5, App. G |

| | | | |
|---|---|---|---|
| **RESULTS** | | | |
| Study selection | 17 | Give numbers of studies screened, assessed for eligibility, and included in the review, with reasons for exclusions at each stage, ideally with a flow diagram. | 6, 9-10, App. J |
| Study characteristics | 18 | For each study, present characteristics for which data were extracted (e.g., study size, PICOS, follow-up period) and provide the citations. | 7-9, App. F, H |
| Risk of bias within studies | 19 | Present data on risk of bias of each study and, if available, any outcome level assessment (see item 12). | App. I |
| Results of individual studies | 20 | For all outcomes considered (benefits or harms), present, for each study: (a) simple summary data for each intervention group (b) effect estimates and confidence intervals, ideally with a forest plot. | 14-16 |
| Synthesis of results | 21 | Present results of each meta-analysis done, including confidence intervals and measures of consistency. | 11-13 |
| Risk of bias across studies | 22 | Present results of any assessment of risk of bias across studies (see Item 15). | App. K |
| Additional analysis | 23 | Give results of additional analyses, if done (e.g., sensitivity or subgroup analyses, meta-regression [see Item 16]). | App. L |
| **DISCUSSION** | | | |
| Summary of evidence | 24 | Summarize the main findings including the strength of evidence for each main outcome; consider their relevance to key groups. | 9-10 |
| Limitations | 25 | Discuss limitations at study and outcome level (e.g., risk of bias), and at review-level (e.g., incomplete retrieval of identified research, reporting bias). | 18 |
| Conclusions | 26 | Provide a general interpretation of the results in the context of other evidence, and implications for future research. | 17-18 |
| **FUNDING** | | | |
| Funding | 27 | Describe sources of funding for the systematic review and other support (e.g., supply of data); role of funders for the systematic review. | Title Page |

**Supplementary Appendix B: Meta-analysis search procedure**

To perform the present meta-analysis, we collected published papers and preprints regarding the seroprevalence and/or infection fatality rate of COVID-19. To identify these studies, we systematically performed online searches in MedRxiv, Medline, PubMed, and Google Scholar using the criterion *((“infection fatality rate” or “IFR” or “seroprevalence” or “antibodies”) and (“COVID-19” or “SARS-Cov-2”))*. We also used a search tool created by the University of Zurich for searching EMBASE using the same search criterion.[2] We identified other studies listed in reports by government institutions such as the U.K. Parliament Office.[3] Finally, we confirmed the coverage of our search by referring to two recent meta-analysis studies of the overall IFR for COVID-19, a recent meta-analysis of the ratio of measured seroprevalence to reported cases, and the SeroTracker global dashboard of SARS-CoV-2 seroprevalence studies.[4-7] Our search encompassed studies that were publicly disseminated prior to 25 September 2020. For cases in which a study was identified by the aforementioned search but age-specific seroprevalence was not found, an expanded search was performed to obtain those details using additional keywords (e.g., the location of the study). Data was extracted from studies by three authors and verified prior to inclusion.

**Supplementary Appendix C: Seroprevalence adjustments for test characteristics**

Studies of COVID-19 seroprevalence have utilized a variety of distinct test procedures. Enzyme-linked immunosorbent assays (ELISA) proceed by tagging antibody-antigen interactions with a reporter protein. Chemiluminescent immunoassays (CLA) work similarly by tagging the antigen-antibody interaction with a fluorescent protein. Lateral Flow Assays (LFA), also known as rapid diagnostic tests (RDT), produce a colored band upon antigen-antibody interaction. Microneutralization tests (MNT), recombinant immunofluorescent assays (RIA), and plaque reduction neutralization tests (PRNT) can be performed in laboratories and provide extremely accurate assessments.

Recognizing that SARS-Cov-2 is both novel and hazardous, public regulatory agencies have issued "emergency use authorizations" (EUA) to facilitate the rapid deployment of test kits. Subsequent studies by independent laboratories have reassessed the characteristics of these test kits, in many cases finding markedly different results than those of the manufacturer. Such differences reflect (a) the extent to which test results may be affected by seemingly trivial differences in its implementation, and (b) the extent to which serological properties may vary across different segments of the population. For example, a significant challenge in producing accurate tests is to distinguish COVID-19 antibodies from those associated with other coronaviruses (including the common cold). Consequently, the assessment of test characteristics may vary with seemingly innocuous factors such as the season of the year in which the blood samples were collected.

The reliability of a seroprevalence test depends on its *sensitivity* (the likelihood that the test correctly detects the virus in an infected person) and *specificity* (the likelihood that the test has a negative result for a uninfected person) as well as the true prevalence of the disease. For example, in a population where the true prevalence is low, a test with sensitivity below 100% will yield a substantial fraction of false positive results.

Some seroprevalence studies have reported results for *raw prevalence*, that is, the ratio $P / N$, where $P$ denotes the number of positive test results and $N$ denotes the number of individuals who were tested, with confidence intervals that solely reflect sampling uncertainty (i.e., variations associated with drawing a finite sample of $N$ observations). However, such estimates and confidence intervals may be misleading in the absence of adjustments that reflect the sensivity and specificity of the test method (except in the case of "gold standard" methods with 100% specificity and 100% sensitivity). Indeed, a recent systematic review and meta-analysis found very substantial divergences in sensitivity and specificity of COVID-19 serological tests.[8]

If the sensitivity and specificity of the test are assumed to be known with certainty, then the *test-adjusted prevalence* can be computed using the Gladen-Rogan formula[9] as follows:

$$adjusted\ prevalence\ =\ \frac{raw\ prevalence\ +\ specificity - 1}{sensitivity\ + specificity - 1}$$

Likewise, the test-adjusted confidence interval can be computed by multiplying the raw confidence interval by the factor 1/(*sensitivity* + *specificity* – 1).

Some recent seroprevalence studies have reported test-adjusted results using the Gladen-Rogan formula.[10-15] However, a number of other seroprevalence studies have only reported raw prevalence results.[12, 16-20] To facilitate the consistency of our meta-analysis, we follow the same approach of adjusting those raw prevalence results using the Gladen-Rogan formula, as reported in the table below.

Nonetheless, the sensitivity and specificity of COVID-19 antibody test kits should generally not be treated as parameters known with certainty. Indeed, the U.S. Food and Drug Administration reports 95% confidence intervals for each of these test properties in its information sheet for all EUA test kits.[21] In particular, following the approach of Gelman and Carpenter (2020), we use a Bayesian procedure to compute seroprevalence estimates and confidence intervals that incorporate uncertainty about the sensitivity and specificity of the test kit used in each of these seroprevalence studies.[22, 23] As shown in Table C1, the contrast between the Gladen-Rogan vs. Bayesian results are most striking for study observations obtained using a relatively small sample size in a context of low prevalence. In some cases, the Bayesian confidence interval indicates that the level of seroprevalence cannot be distinguished from zero, and hence those observations are excluded from our metaregression (see Appendix X).

### Table C1: Raw vs. test-adjusted measures of seroprevalence

| Location | Age Group | Sample (N) | Prevalence (%) Raw | Prevalence (%) Rogan-Gladen | Prevalence (%) Bayesian | 95% CI Lower Bound Raw | 95% CI Lower Bound Rogan-Gladen | 95% CI Lower Bound Bayesian | 95% CI Upper Bound Raw | 95% CI Upper Bound Rogan-Gladen | 95% CI Upper Bound Bayesian |
|---|---|---|---|---|---|---|---|---|---|---|---|
| Belgium[24] | 0-24 | 1263 | 6.0 | 6.7 | 5.7 | 4.2 | 4.7 | 2.0 | 8.6 | 9.6 | 8.4 |
| | 25-44 | 1710 | 5.9 | 6.6 | 5.4 | 4.2 | 4.7 | 1.9 | 8.3 | 9.2 | 8.0 |
| | 45-64 | 1831 | 6.2 | 6.9 | 5.6 | 4.7 | 5.2 | 2.1 | 8.3 | 9.2 | 8.3 |
| | 65-74 | 878 | 4.1 | 4.6 | 3.6 | 2.3 | 2.6 | 0.7 | 7.2 | 8.0 | 6.1 |
| | 75-84 | 816 | 7.0 | 7.8 | 7.0 | 4.2 | 4.7 | 3.4 | 11.7 | 13.0 | 10.1 |
| | 85+ | 809 | 13.2 | 14.7 | 14.2 | 8.9 | 9.9 | 9.8 | 19.6 | 21.8 | 18.7 |
| France[25] | 0–39 | 2262 | 10.8 | 12.0 | NA | 9.6 | 10.6 | NA | 12.1 | 13.5 | NA |
| | 40–49 | 2897 | 11.5 | 12.7 | NA | 10.3 | 11.4 | NA | 12.6 | 14.0 | NA |
| | 50–59 | 3019 | 5.8 | 6.5 | NA | 5.0 | 5.5 | NA | 6.7 | 7.4 | NA |
| | 60–69 | 3272 | 4.1 | 4.5 | NA | 3.4 | 3.8 | NA | 4.7 | 5.3 | NA |
| | 70+ | 3175 | 3.1 | 3.4 | NA | 2.5 | 2.7 | NA | 3.7 | 4.1 | NA |
| Hungary[17] | 14-39 | 3353 | 0.5 | 0.1 | 0.2 | 0.3 | 0.0 | 0.0 | 0.9 | 0.5 | 0.6 |
| | 40-64 | 4735 | 0.7 | 0.3 | 0.3 | 0.4 | 0.0 | 0.0 | 1.0 | 0.6 | 0.7 |
| | 65+ | 2386 | 0.8 | 0.4 | 0.4 | 0.4 | 0.0 | 0.1 | 1.3 | 0.9 | 0.9 |
| Ireland[26] | 12–29 | 519 | 1·9 | 1·5 | NA | 0·7 | 0·3 | NA | 3·1 | 2·7 | NA |
| | 30–49 | 746 | 1·6 | 1·2 | NA | 0·7 | 0·3 | NA | 2·5 | 2·1 | NA |
| | 50–69 | 686 | 1·6 | 1·2 | NA | 0·7 | 0·3 | NA | 2·5 | 2·2 | NA |
| Italy[16] | 0-19 | 9460 | 2.2 | 1.8 | 1.8 | 1.7 | 1.3 | 1.2 | 2.8 | 2.4 | 2.2 |
| | 20-29 | 11302 | 2.1 | 1.7 | 1.7 | 1.7 | 1.3 | 1.1 | 2.4 | 2.0 | 2.1 |
| | 30-49 | 13816 | 2.4 | 2.0 | 2.0 | 2.1 | 1.7 | 1.4 | 2.8 | 2.4 | 2.4 |
| | 50-59 | 10639 | 3.1 | 2.7 | 2.7 | 2.7 | 2.3 | 2.1 | 3.5 | 3.1 | 3.2 |
| | 60-69 | 8324 | 2.6 | 2.2 | 2.2 | 2.1 | 1.7 | 1.6 | 2.9 | 2.5 | 2.7 |
| | 70+ | 11118 | 2.5 | 2.1 | 2.1 | 2.1 | 1.7 | 1.5 | 2.9 | 2.5 | 2.5 |
| Portugal[18] | 0-9 | 404 | 2.0 | 2.2 | 1.8 | 0.8 | 0.9 | 0.1 | 5.4 | 6.0 | 3.9 |
| | 10-19 | 377 | 2.1 | 2.4 | 1.8 | 0.8 | 0.9 | 0.1 | 5.5 | 6.1 | 4.2 |
| | 20-39 | 377 | 0.8 | 0.9 | 0.8 | 0.1 | 0.1 | 0.1 | 4.8 | 5.3 | 2.3 |
| | 40-59 | 479 | 2.3 | 2.6 | 2.0 | 0.9 | 1.0 | 0.2 | 5.9 | 6.6 | 4.2 |
| | 60+ | 664 | 2.4 | 2.7 | 2.0 | 1.1 | 1.2 | 0.2 | 4.9 | 5.4 | 4.0 |

#### Table C1: Raw vs. test-adjusted measures of seroprevalence *(contd.)*

| | | | | | | | | | | | |
|---|---|---|---|---|---|---|---|---|---|---|---|
| Spain[20] | 0-9 | 1590 | 2.8 | 2.4 | 2.4 | 1.5 | 1.1 | 1.5 | 5.2 | 4.8 | 3.4 |
| | 10-19 | 4937 | 3.3 | 2.9 | 2.9 | 2.4 | 2.0 | 2.3 | 4.5 | 4.1 | 3.5 |
| | 20-29 | 4808 | 3.8 | 3.4 | 3.4 | 2.8 | 2.4 | 2.7 | 5.2 | 4.8 | 4.1 |
| | 30–39 | 6445 | 3.5 | 3.1 | 3.0 | 2.6 | 2.2 | 2.4 | 4.5 | 4.1 | 3.6 |
| | 40–49 | 9670 | 4.0 | 3.6 | 3.6 | 3.1 | 2.8 | 3.0 | 4.9 | 4.5 | 4.1 |
| | 50–59 | 9652 | 4.1 | 3.7 | 3.7 | 3.2 | 2.8 | 3.1 | 5.1 | 4.7 | 4.2 |
| | 60–69 | 7564 | 3.8 | 3.4 | 3.4 | 2.9 | 2.5 | 2.8 | 4.9 | 4.5 | 3.9 |
| | 70+ | 7293 | 3.5 | 3.1 | 3.1 | 2.3 | 1.9 | 2.5 | 5.4 | 5.0 | 3.6 |
| Indiana, USA[19] | 0-39 | 1011 | 1.4 | 1.0 | 1.0 | 0.7 | 0.3 | 0.2 | 2.2 | 1.8 | 1.9 |
| | 40-59 | 1301 | 1.1 | 0.7 | 0.7 | 0.5 | 0.1 | 0.1 | 1.8 | 1.4 | 1.4 |
| | 60+ | 1206 | 0.7 | 0.3 | 0.4 | 0.3 | 0.0 | 0.0 | 1.3 | 0.9 | 1.1 |

**Supplementary Appendix D: Countries with comprehensive tracing programs**

| Country | Cases *(thousands)* | Tests per confirmed case |
|---|---|---|
| New Zealand | 1·1 | 1862 |
| Australia | 6·7 | 1054 |
| South Korea | 10·8 | 576 |
| Lithuania | 1·4 | 415 |
| Iceland | 1·8 | 321 |
| Slovakia | 1·4 | 194 |
| Latvia | 0·8 | 191 |
| Austria | 15·4 | 115 |
| Slovenia | 1·4 | 112 |
| Czech Republic | 7·6 | 104 |
| Greece | 2·6 | 95 |
| Denmark | 9·0 | 94 |
| Estonia | 1·7 | 70 |
| Luxembourg | 3·8 | 68 |
| Israel | 15·8 | 57 |
| Norway | 7·7 | 47 |
| Poland | 14·0 | 37 |
| Hungary | 2·8 | 36 |
| Portugal | 24·7 | 35 |
| Belgium | 49·9 | 32 |
| Germany | 159·1 | 31 |
| Finland | 4·9 | 31 |
| Switzerland | 29·3 | 27 |
| Spain | 215·2 | 27 |
| Japan | 14·1 | 25 |
| Italy | 203·6 | 19 |
| Colombia | 6·2 | 16 |
| Canada | 51·6 | 15 |
| Ireland | 20·3 | 13 |
| Turkey | 117·6 | 13 |
| Chile | 14·9 | 13 |
| United Kingdom | 167·2 | 10 |
| Netherlands | 38·8 | 8 |
| United States | 1039·9 | 8 |
| Mexico | 17·8 | 4 |
| France | 129·6 | NA |
| Sweden | 21·7 | NA |

*Note*: This table reports data for all OECD countries as of 30 April except Lithuania (28 April) and Poland (5 May); data on tests per confirmed case was not available for France and Sweden.[27] A national seroprevalence study of the Czech Republic found that infections exceeded confirmed cases by a factor of 5, suggesting that comprehensive tracing requires substantially more than 100 tests per confirmed case. By contrast, a seroprevalence study of Iceland indicates that its tracing program was effective in identifying a high proportion of SARS-CoV-2 infections.[28] For example, the Korea Center for Disease Control analyzed a total of 2995 serum samples collected for the National Health and Nutrition Examination Survey between 21 April and 13 August, and only one of those specimens tested positive for antibodies and neutralizing antibodies.[29] Similarly, a seroprevalence study of 1500 outpatients in Seoul found only one positive result.[30]

**Supplementary Appendix E: Prevalence vs reported cases in Iceland**

| Age Group | Reported Cases | Estimated Infections | *Confidence Interval* | | Ratio of Infections to Reported Cases | *Confidence Interval* | |
|---|---|---|---|---|---|---|---|
| | | | *Lower* | *Upper* | | *Lower* | *Upper* |
| *30-39* | 289 | 469 | *469* | *703* | 1·6 | *1·6* | *2·4* |
| *40-49* | 357 | 644 | *473* | *859* | 1·8 | *1·3* | *2·4* |
| *50-59* | 306 | 337 | *211* | *547* | 1·1 | *0·7* | *1·8* |
| *60-69* | 213 | 225 | *188* | *375* | 1·1 | *0·9* | *1·8* |
| *70-79* | 63 | 70 | *63* | *304* | 1·1 | *1·0* | *4·8* |
| *80+* | 25 | 26 | *13* | *319* | 1·0 | *0·5* | *12·8* |
| *All 30+* | 1253 | 1771 | *1415* | *3109* | 1·41 | *1·13* | *2·48* |

*Sources*: Cases are reported by Iceland Directorate of Health as of 14 June 2020, when Iceland had 1796 recovered cases, 10 fatalities, and 4 individuals in isolation (none hospitalized).[31] Estimated infections and 95% confidence intervals are taken from the prevalence study of Gudbjartsson et al. (2020), which conducted tests of a random sample of the general population on 16–31 March 2020.[32] As of 21 April 2020 (three weeks after the conclusion of that study), there were 1785 reported cases (98·6% of the total reported cases as of 14 June 2020).

#### Supplementary Appendix F: Age-specific fatality data and source information

##### F.1: Fatality data for European seroprevalence studies with representative samples

| Location and source of fatality data | Study midpoint date in 2020 | Fatality reporting date in 2020 | Age group, years | Fatalities |
|---|---|---|---|---|
| Belgium[33] | April 23 | May 16 | 0–24 | 1 |
| | | | 25–44 | 30 |
| | | | 45–64 | 409 |
| | | | 65–74 | 1061 |
| | | | 75–84 | 2144 |
| | | | 85+ | 5087 |
| England[34] | July 1 | July 29 | 0–17 | 11 |
| | | | 18–24 | 30 |
| | | | 25–34 | 131 |
| | | | 35–44 | 394 |
| | | | 45–54 | 1348 |
| | | | 55–64 | 3605 |
| | | | 65–74 | 7631 |
| | | | 75+ | 38629 |
| France[35-38] | May 14 | May 31 | 0–39 | 78 |
| | | | 40–49 | 167 |
| | | | 50–59 | 610 |
| | | | 60–69 | 1494 |
| | | | 70+ | 15050 |
| Geneva, Switzerland[39] | April 26 | June 1 | 0–19 | 0 |
| | | | 20–49 | 2 |
| | | | 50–64 | 16 |
| | | | 65+ | 268 |
| Ireland[40] | July 4 | August 2 | 15–44 | 19 |
| | | | 45–64 | 98 |
| Italy[34] | July 16 | August 13 | 0–19 | 4 |
| | | | 20–29 | 16 |
| | | | 30–49 | 369 |
| | | | 50–59 | 1186 |
| | | | 60–69 | 3433 |
| | | | 70+ | 29134 |
| Netherlands[34] | April 9 | May 7 | 0–49 | 40 |
| | | | 50–59 | 137 |
| | | | 60–69 | 454 |
| | | | 70–79 | 1539 |
| | | | 80+ | 2426 |
| Portugal[34] | June 14 | July 12 | 0–9 | 0 |
| | | | 10–19 | 0 |
| | | | 20–39 | 4 |
| | | | 40–59 | 75 |
| | | | 60+ | 1581 |
| Spain[41] | May 25 | July 15 | 0–9 | 5 |
| | | | 10–19 | 6 |
| | | | 20–29 | 35 |
| | | | 30–39 | 77 |
| | | | 40–49 | 295 |
| | | | 50–59 | 1023 |
| | | | 60–69 | 3049 |
| | | | 70+ | 24647 |

*Notes*: (1) The France seroprevalence study encompassed the regions of Grand Est, Ile de France, and Nouvelle Aquitaine. Total confirmed COVID-19 fatalities (including deaths in hospitals and in other medical and social institutions) were reported by region as of May 31.[35] The age composition of hospital fatalities for each region (published on June 3, May 28, and May 28, respectively) was used to compute total age-specific fatalities as of May 31.[36-38] Fatalities in other medical and social institutions were allocated to the 70+ age group. (2) Spain had 29137 laboratory-confirmed COVID-19 fatalities as of 15 July 2020, including 9909 deaths in nursing homes. Age-specific fatalities outside of nursing homes are reported by Pastor-Barriuso et al. (2020).[41] Abellán et al. (2020) estimated that 4% of Spanish nursing home residents were aged 65-69 while the remaining 96% were aged 70+.[42] We used those proportions to allocate the 9909 nursing home deaths to the corresponding age groups.

**F.2: Fatality data for European and Canadian seroprevalence studies with convenience samples**

| Location and source of fatality data | Study midpoint date in 2020 | Fatality reporting date in 2020 | Age group, years | Fatalities |
|---|---|---|---|---|
| France[43, 44] | April 12 | May 31 | 0–9 | 3 |
| | | | 10–19 | 3 |
| | | | 20–29 | 21 |
| | | | 30–39 | 84 |
| | | | 40–49 | 231 |
| | | | 50–59 | 860 |
| | | | 60–69 | 2204 |
| | | | 70–79 | 5650 |
| | | | 80+ | 19746 |
| Ontario[45] | June 17 | July 15 | 0–19 | 1 |
| | | | 20–59 | 122 |
| | | | 60+ | 2600 |
| Sweden[46] | May 10 | June 18 | 0–19 | 1 |
| | | | 20–49 | 63 |
| | | | 50–69 | 504 |
| | | | 70+ | 4485 |

*Notes*: France had a total of 28802 confirmed COVID-19 fatalities as of 31 May 2020, including 18475 deaths in hospitals and 10327 deaths in other medical and social institutions; France does not have real-time reporting of deaths in personal homes.[44] Age-specific COVID-19 fatalities are only reported for hospitals; therefore, age-specific COVID-19 fatalities for other medical and social institutions are allocated proportionally for ages 70–79 and 80+ based on the relative incidence of overall excess mortality during weeks 10 to 22 for those two age groups as reported by Eurostat (which collects that data daily from France Santé Publique).[43]

##### F.3 Fatality data for U.S. seroprevalence studies

| Location and source of fatality data | Study midpoint date in 2020 | Fatality reporting date in 2020 | Age Group | Fatalities |
|---|---|---|---|---|
| Atlanta[47] | May 1 | May 31 | 0–17 | 1 |
| | | | 18–49 | 20 |
| | | | 50–64 | 51 |
| | | | 65+ | 294 |
| Connecticut[48] | April 30 | May 28 | 0–19 | 2 |
| | | | 20–49 | 75 |
| | | | 50–59 | 157 |
| | | | 60+ | 3633 |
| Indiana[49] | April 27 | May 25 | 0–39 | 20 |
| | | | 40–59 | 148 |
| | | | 60+ | 1864 |
| Louisiana[50] | April 5 | May 6 | 0–18 | 1 |
| | | | 19–49 | 85 |
| | | | 50–59 | 126 |
| | | | 60+ | 1053 |
| Miami[51] | April 8 | May 6 | 0–18 | 0 |
| | | | 19–49 | 61 |
| | | | 50–59 | 169 |
| | | | 60+ | 1060 |
| Minneapolis[52] | May 5 | June 4 | 0–18 | 0 |
| | | | 19–49 | 18 |
| | | | 50–59 | 47 |
| | | | 60+ | 928 |
| Missouri[53] | April 23 | May 23 | 0–19 | 0 |
| | | | 20–49 | 18 |
| | | | 50–59 | 43 |
| | | | 60+ | 620 |
| New York[54] | April 23 | May 21 | 0–19 | 12 |
| | | | 20–39 | 482 |
| | | | 40–49 | 1026 |
| | | | 50–59 | 2764 |
| | | | 60+ | 24376 |
| Philadelphia[55, 56] | April 19 | May 17 | 0–18 | 1 |
| | | | 19–49 | 57 |
| | | | 50–59 | 323 |
| | | | 60+ | 2639 |
| Salt Lake City[57] | May 22 | June 19 | 0–44 | 4 |
| | | | 45–64 | 31 |
| | | | 65+ | 90 |
| San Francisco[58] | April 25 | May 25 | 0–18 | 0 |
| | | | 19–49 | 25 |
| | | | 50–59 | 66 |
| | | | 60+ | 333 |
| Seattle[59] | March 27 | April 26 | 0–19 | 0 |
| | | | 20–39 | 8 |
| | | | 40–59 | 69 |
| | | | 60+ | 700 |

Note: Some seroprevalence age brackets were adjusted (+/- 5 years) to match the age structure of that location's COVID-19 fatality report. For Pennsylvania and Utah, county-level data as of May 17 is available for total fatalities but not reported by age group; consequently, the statewide age distribution of fatalities was used to allocate county-level fatalities by age for each of those locations.

**F.4 Fatality data for comprehensive tracing countries**

| Location and source of fatality data | Reporting date | Age group, years | Fatalities |
|---|---|---|---|
| Australia[60] | June 12 | 0–39 | 0 |
| | | 40–59 | 3 |
| | | 60–69 | 13 |
| | | 70–79 | 31 |
| | | 80+ | 55 |
| Iceland[31] | June 14 | 0–29 | 0 |
| | | 30–59 | 1 |
| | | 60–69 | 2 |
| | | 70–79 | 3 |
| | | 80+ | 4 |
| Korea[61] | July 11 | 0–29 | 0 |
| | | 30–39 | 2 |
| | | 40–49 | 3 |
| | | 50–59 | 15 |
| | | 60–69 | 41 |
| | | 70–79 | 84 |
| | | 80+ | 144 |
| Lithuania[62] | June 21 | 0–39 | 0 |
| | | 40–49 | 1 |
| | | 50–59 | 3 |
| | | 60–69 | 12 |
| | | 70–79 | 23 |
| | | 80+ | 37 |
| New Zealand[63] | July 9 | 0–59 | 0 |
| | | 60–69 | 3 |
| | | 70–79 | 7 |
| | | 80+ | 12 |

Note: Age-specific fatality data for Lithuania was published as of 01 June 2020, at which point there was a total of 70 reported fatalities; thus, the six subsequent fatalities through 22 June 2020 were assumed to have the same age distribution as the fatalities through 01 June 2020.

**Supplementary Appendix G: Metaregression methodology**

To analyze IFR by age, we use meta-regression with random effects, using the *meta regress* procedure in *Stata* v16.[64, 65] We used a random-effects procedures to allow for residual heterogeneity between studies and across age groups by assuming that these divergences are drawn from a Gaussian distribution. The procedure provides reasonable results even if the errors are not strictly normal but may be unsatisfactory if the sample includes large outliers or the distribution of groups is not unimodal. In analytical terms, this framework can be expressed as follows:

$$log\left(IFR_{ij}\right) = \alpha \ + \ \beta * age_{ij} \ + \ \epsilon_{ij} \ + \ u_{ij}$$

$$\text{where } u_{ij} \sim N(0, \tau^2) \text{ and } \epsilon_{ij} \sim N\left(0, \sigma_{ij}^2\right)$$

In this specification, $IFR_{ij}$ is the estimated IFR in study *i* for age group *j*, $age_{ij}$ denotes the median age of that group, $\epsilon_{ij}$ denotes the source of idiosyncratic variations for that particular location and age group, and $u_{ij}$ denotes the random effects that characterize any systematic deviations in outcomes across locations and age groups. Under the maintained assumption that each idiosyncratic term $\epsilon_{ij}$ has a normal distribution, the idiosyncratic variance is $\sigma_{ij}^2 = ((U_{ij} - L_{ij})/3.96)^2$, where $U_{ij}$ and $L_{ij}$ denote the upper and lower bounds of the 95% confidence interval for that study-age group. The random effects $u_{ij}$ are assumed to be drawn from a homogeneous distribution with zero mean and variance $\tau^2$. The null hypothesis of $\tau^2 = 0$ characterizes the case in which there are no systematic deviations across studies or age groups. If that null hypothesis is rejected, then the estimated value of $\tau^2$ encapsulates the magnitude of those systematic deviations.

Under our baseline specification, the infection fatality rate increases exponentially with age—a pattern that has been evident in prior studies of age-specific case fatality rates.[66, 67] Consequently, our meta-regression is specified in logarithmic terms, with the slope coefficient $\beta$ encapsulating the impact of higher age on *log(IFR)*. Consequently, the null hypothesis that IFR is unrelated to age can be evaluated by testing whether the value of $\beta$ is significantly different from zero. If that null hypothesis is rejected, then the estimated values of $\alpha$ and $\beta$ characterize the estimated relationship between log(IFR) and age. Consequently, the predicted relationship between IFR and age can be expressed as follows:

$$IFR \ = \ e^{\alpha \ + \ \beta * age_{ij}}$$

The 95% confidence interval for this prediction can obtained using the delta method. In particular, let $IFR_a$ denote the infection fatality rate for age $a$, and let $\sigma_c$ denote the standard error of the meta-regression estimate of log($IFR_a$). If $IFR_a$ has a non-zero value, then the delta method indicates that its standard error equals $\sigma_c$ / $IFR_a$ , and this standard error is used to construct the confidence interval for $IFR_a$ at each age $a$. Likewise, the prediction interval for log($IFR_a$) is computed using a standard error of $\sigma_c + \ \tau$ that incorporates the systematic variation in the random effects across studies and age groups, and hence the corresponding prediction interval for $IFR_a$ is computed using a standard error of $(\sigma_c + \ \tau)/IFR_a$ .

In estimating this metaregression, we exclude observations for which the lower bound of the 95% confidence interval for seroprevalence of that particular age group equals zero, and hence the upper bound of that age-specific IFR is not well defined. Similarly, we exclude observations for which the lower bound of the 95% confidence interval for seroprevalence is less than the observed COVID-19 mortality rate for that age group, since such observations would imply an upper bound for the IFR that exceeds 100%. Finally, we exclude observations for which no COVID-19 fatalities were recorded for a given age group and hence the implied value of the infection fatality rate is at its lower bound of zero and the corresponding confidence interval cannot be precisely determined.

#### Supplementary Appendix H: List of excluded studies

##### H.1 Studies excluded due to absence of age-specific prevalence or fatality data

| Location | Description |
|---|---|
| Alberta, Canada[68] | On July 30, Alberta's chief medical officer announced the results of a serology study of 9400 blood specimens collected for other purposes and indicated that "less than one percent" were positive. No further details were available as of September 17, 2020. |
| Ariano Irpino, Italy[69] | This seroprevalence study collected specimens in late May from 13444 individuals (about 75% of municipality residents) and found a raw prevalence of 4·83%. No age-specific results were reported. |
| Australia[70] | This study analyzed specimens from 2991 individuals undergoing elective surgery during May–June 2020 and found test-adjusted seroprevalence of 0·28% (CI: 0–0·71%). No age-specific seroprevalence results were reported. Although the sample of patients may not be fully representative of the Australian population, these results confirm that prevalence in Australia was indistinguishable from zero at the time of the study, as expected for a comprehensive tracing program (see Supplementary Appendix C). |
| Austria[71] | Statistik Austria conducted an experimental study in which specimens were collected from 269 individuals ages 16+. The test-adjusted seroprevalence was 4·71% (CI: 1·36–7·97%). No age-specific results were reported. |
| Bad Feilnbach, Germany[72] | This study collected specimens from a random sample of 2153 adults between 23 June and 4 July 2020 and found raw seroprevalence of 6·0%. No age-specific results were reported as of September 17. |
| Baton Rouge, Louisiana, USA[73] | This study analyzed specimens from a random sample of 2138 individuals between July 15–31 and found seroprevalence of 3·6%. No age-specific seroprevalence results were reported. |
| Blaine County, Idaho, USA[74] | This study collected specimens from 972 individuals on May 4-19 and found an IgG prevalence of 22·7% (CI: 20–25·5%). The authors concluded that "the small number of county deaths (*n*=5) makes estimating the infection fatality rate unreliable." No age-specific fatality data is publicly available for this county. |
| Bolinas, California, USA[75] | This study collected specimens from a random sample of 1312 confirmed residents of Bolinas on April 20-24. The test-adjusted seroprevalence was 0·16% (CI: 0·02–0·46%). No age-specific seroprevalence results were reported. |
| Bonn, Germany[76] | This study analyzed specimens from 4771 individuals participating in the Rhineland Study, an ongoing community-based prospective cohort study of Bonn residents ages 30 years and older. Specimens were collected between 24 April and 30 June 2020 and analyzed using a combination of ELISA, RIA, and PRNT methods. The ELISA assay found 46 positive results and indicated seroprevalence of 0·97% (CI 0·72–1·30%). By contrast, the PRNT assay found only 17 positive results and indicated a markedly lower seroprevalence of 0·36% (CI: 0·21–0·61%). No age-specific seroprevalence results were reported. |
| British Columbia, Canada[77] | This study analyzed 885 laboratory specimens from outpatient clinics for the period May 15-27 and found only four positive cases (0·6%). No age-specific seroprevalence was reported. |
| Burlington, Vermont, USA[78] | This study analyzed specimens from 454 primary care patients at a Level 1 medical center in Burlington on 25–28 June 2020 and found 10 positive results using a two-step serologic assay and found raw prevalence of 2·2 percent (CI: 0·8–3·6%). The median age of individuals with positive vs. negative results were nearly identical (51·9 vs. 51·4 years). |
| Caldari Ortona, Italy[79] | This study collected specimens from 640 residents on April 18-19 and found raw prevalence of 12%. No age-specific results were reported. |
| Chelsea, Massachusetts, USA[80] | This study collected specimens from 200 pedestrians and found a raw seroprevalence of 22·5% using an IgG LFA. No age-specific results were reported. |

| | |
|---|---|
| Connecticut, USA[81] | This study analyzed specimens from a random sample of 505 adults residing in non-congregate settings. The sample design reflected the assumption of statewide prevalence of 10% (roughly similar to that of the neighboring state of New York) with the aim of obtaining prevalence estimates with precision of 2% at a confidence level of 90%. However, the study obtained a much lower estimated prevalence of 3·1% (95% CI: 1·1–5·1%). Consequently, the sample size proved to be insufficient to provide reliable age-specific results; the margin of error exceeds the estimated prevalence for all age groups reported in the study. |
| Czech Republic[82] | The Czech Ministry of Health conducted a large-scale seroprevalence survey on April 23-May 1, collecting specimens from a random sample of 22316 residents and testing for IgG antibodies using the Wantai test kit. Only 107 positive cases were identified (raw prevalence = 0·4%), and hence the test-adjusted confidence intervals include the lower bound of zero prevalence. That result is consistent with the very low number of reported cases in the Czech Republic as of early May; for example, Prague had only 1,638 reported cases for a population of 1·3 million. |
| Denmark[83] | This study analyzed specimens from a random sample of 2427 individuals in early June and identified 34 positive cases, yielding a test-adjusted prevalence of 1·2% (CI: 0·7–1·7%). Age-specific estimates were not reported as of September 17. |
| Faroe Islands Denmark[84] | This study analyzed specimens from a random sample of 1075 participants during late April and obtained 6 positive results; the test-adjusted prevalence was 0·7% (CI: 0·3–1·3%). No age-specific results were reported. |
| Finland[85] | Finland National Institute for Health and Welfare has been conducting an ongoing study of seroprevalence using random sampling of the population. Each specimen is initially screened for antibodies using a rapid test, and all specimens with positive screening results are analyzed using a microneutralization test (MNT) with confirmed specificity of 100%. As of August 8, this process screened 3155 specimens and obtained 8 positive MNT results (0.25%). No age-specific results were reported as of September 17. |
| Gangelt, Germany[86] | This study analyzed specimens from a random sample of 919 participants from the municipality of Gangelt (population 12,597) on March 31 to April 6 and obtained a test-adjusted prevalence of 15·5% (CI: 12·3–19·0%). Official government reports indicate that Gangelt had 7 COVID-19 fatalities at the time of the study but the death toll rose to 12 by late June, indicating an overall IFR of about 0·6%, similar to the IFR for Geneva. Age-specific fatalities have not been reported for Gangelt. |
| Greece[87] | This study analyzed residual serum specimens from 6586 individuals collected during March and April and found 24 positive results. The test-adjusted prevalence was 0% (CI: 0–0·23%). Prevalence was reported for four age groups (0–29, 30–49, 50–69, and 70+); each of those confidence intervals included the lower limit of 0%. |
| Hibino, Japan[88] | This study analyzed specimens from 615 employees of a Japanese company in Tokyo, including 350 individuals who were tested on two separate occasions. Among the 95 individuals with positive IgG results on the first occasion, there were 12 with negative IgG results in their second test. Among the 252 with an initial negative result, 54 had positive IgG results in their second test. No age-specific test results were reported. |
| Hideaway, Texas USA[89] | This study analyzed specimens from 293 adults and found 7 positive results. Age-specific results were not reported. |
| Ischgl, Austria[90] | This study analyzed specimens from 184 adults in Ischgl (an Austrian municipality of 1,604 residents) and obtained 85 positive results, i.e., prevalence of 46·2%. The study reported the fraction of positive results for specific age groups (4 out of 11 adults 55-64 years, 2 out of 8 adults 65-74 years, and 1 out of 2 adults ages 75+) but did not report test-adjusted estimates or confidence intervals by age group. Ischgl had only 2 reported COVID-19 fatalities as of July 1. |
| Israel[91] | Israel Health Ministry initiated a large-scale seroprevalence study in May. Subsequent media reports indicated that initial tests of 70000 Israelis indicated that prevalence varied significantly across regions and health organizations. No age-specific results had been released as of September 17. |
| Japanese Evacuees[92] | This study performed PCR tests on 565 Japanese citizens expatriated from Wuhan, China. There were eight positive tests, indicating a raw prevalence of 1·4%, but assessment of age-specific prevalence or IFRs is not feasible given the small sample, low prevalence, and lack of data on case outcomes. |
| Jersey, United Kingdom[93] | This study collected samples from 629 households comprising 1,062 individuals and estimated seroprevalence at 4·2% (CI 2·9 to 5·5%), indicating that about 3,300 Jersey residents have been infected. Jersey has had 30 COVID-19 fatalities (as of July 15), and hence the overall IFR is about 1% (similar to that of NYC). However, the seroprevalence sample is too small to facilitate accurate assessments of age-specific IFRs; for ages 55+, there were 258 samples and 12 positive cases, |

| | |
|---|---|
| Lisbon, Portugal[94] | This study analyzed specimens from 300 hospital patients and healthcare workers, 2500 university staff, and 187 adult volunteers and found that the ability to detect SARS-CoV-2 antibodies remained robust for five months in a large proportion of previously virus-positive screened subjects. |
| Louisville, Kentucky, USA[95] | This study analyzed specimens from 2237 individuals, including 509 who responded to mailed invitations and 1728 who volunteered after hearing about the study via news or social media, and found raw seroprevalence of 4·1% (CI: 3·2–5·1%). Age-specific results were not reported as of September 17. |
| Miami-Dade County, Florida, USA[96] | This study analyzed samples from 2,357 individuals in April and obtained 65 positive IgG results; an additional 275 individuals were tested in June with 4 positive results. Test-adjusted seroprevalence estimates and confidence intervals have not been published as of September 17. |
| New York City, New York, USA[97] | This study analyzed seroprevalence using specimens from four groups of patients (Cardiology, OB/GYN, Oncology, and Surgery) starting in mid-February. For the final week of the study (April 19), positive results were obtained for 47 of 243 patients; that seroprevalence estimate of 19·3% is well-aligned with the results of the New York Department of Health study. However, the sample size of this cohort is too small for assessing age-specific IFRs. |
| Neustadt-am-Rennsteig, Germany[98] | This study analyzed seroprevalence of 626 residents (71% of the population of this municipality) and estimated seroprevalence of 8·4% (52 positive cases). However, this sample size is too small for assessing age-specific IFRs. |
| New Orleans, Louisiana, USA[99] | This study analyzed seroprevalence in a random sample of 2,640 participants and obtained a seroprevalence estimate of 6·9% and an IFR of 1·6% (CI 1·5 to 1·7%). The study did not report on age-specific seroprevalence or IFRs. |
| New York City, New York, USA[100] | This study estimated age-specific infection fatality rates in New York City during spring 2020 using case, mortality, and mobility data. No seroprevalence data was utilized. |
| Norbotten, Sweden[101] | This study analyzed a randomly-selected sample of 425 adults and obtained 8 positive results; the test-adjusted seroprevalence was 1·9% (CI: 0·8–3·7%). However, only 2 positive results were for ages 30-64 and 2 positive results for ages 65+, so age-specific prevalence and IFRs cannot be reliably estimated. |
| Norway[102] | Norwegian Institute of Public Health collected 900 residual serum specimens from nine laboratories from various regions of Norway and obtained test-adjusted seroprevalence of 1·0% (CI: 0.1–2·4%). The study found 4 positive results out of 372 specimens for adults ages 25–59 and 2 positive results out of 206 specimens for adults ages 60+. The authors noted that *"these results should be interpreted with caution"* due to the limited size of the sample. |
| Occitania, France[103] | This study analyzed samples from 613 individuals *"exposed to the virus to varying extents mimicking the general population in Occitania"* and found seroprevalence of 1·3% (CI: 0·6–2·6%). The study did not report any age-specific data. |
| Oklahoma, USA[104] | The Oklahoma Department of Health publishes weekly data on raw seroprevalence using samples collected from labs within the state, but its reports do not include test-adjusted estimates, confidence intervals, or age-specific results. |
| Oslo, Norway[105] | As of August 12, this ongoing study had analyzed specimens from 3250 participants in the Norwegian Mother, Father and Child Survey (MoBa) and found seroprevalence of "less than 2 percent." No confidence intervals or age-specific results were reported. |
| Pima County, Arizona, USA[106] | This study analyzed specimens from 5882 self-recruited members of the local community and found 60 specimens with neutralizing antibodies (1·0%). Age-specific seroprevalence was not reported. |
| Rhode Island, USA[107] | This study invited 5000 randomly-selected households, collected samples from "roughly 10 to 15 percent" who agreed to participate, and obtained seroprevalence of 2·2% (CI: 1·1–3·9%). No age-specific results have been reported as of September 17. |
| Riverside County, California, USA[108] | This study tested a randomized sample of 1,726 residents during July and found raw seroprevalence of 5·9%. The press release (issued on July 27) indicated that the results "are still being analyzed"; no test-adjusted seroprevalence results or age-specific findings have been reported as of September 17. |
| San Francisco Mission District, California[109] | This study analyzed active infections and seroprevalence of 3,953 residents in a densely population majority Latinx neighborhood in downtown San Francisco. Positive seroprevalence in older adults was very low (22 out of 3,953) and hence too small for assessing age-specific IFRs. |
| San Miguel County, Colorado, USA[110] | The San Miguel County Health Department assessed seroprevalence in March and April using samples from 5,283 participants (66% of county residents). Raw prevalence was very low (0·53%), with only 3 confirmed positive results for adults ages 60 years and above. |

| | |
|---|---|
| Slovenia[111] | Researchers at the University of Ljubljana assessed seroprevalence using an IgG ELISA test for a random sample of 1,318 participants on April 20 to May 3. Test-adjusted prevalence was 0·9% (CI: 0 to 2·1%), indicating that the sample may have included only 10 infected individuals; no age-specific results were reported. |
| South-East England[112] | This study collected samples from 481 participants of the TwinsUK cohort and obtained 51 positive results (raw prevalence of 12%). No age-specific results were reported. |
| Stockholm, Sweden[113] | This study did not directly assess prevalence but produced estimates of IFR for two age groups (ages 0-69 and 70+) using a novel methodology linking live virus tests, reported cases, and mortality outcomes. The estimated IFR was 4·3% for ages 70+. |
| Stockholm Districts, Sweden[114] | This study analyzed samples from 213 randomly selected individuals in two residential areas of Stockholm on June 17–18 and found markedly different seroprevalence rates of 4·1% and 30%, respectively. No age-specific results were reported. |
| Stockholm Region, Sweden[115] | Stockholm County began offering antibody testing on a free walk-up basis. As of July 20, 166,431 antibody tests had been performed, of which 17·7% were positive. No demographic data or test-adjusted seroprevalence results had been reported as of September 17. |
| Miyagi, Osaka, and Tokyo, Japan[116] | This study collected samples from randomly-selected residents of three cities on June 1-7 and used two IgG test kits (Abbott and Roche); results were deemed “positive” only if confirmed by both tests Estimated seroprevalence was 0·1% in Tokyo (2 positive results from 1,971 specimens), 0·17% in Osaka (5 positive results from 2,970 specimens), and 0·03% in Miyagi (1 positive result from 3,009 specimens). No age-specific prevalence estimates were reported. |
| United States[117] | Seroprevalence estimates are reported in the U.S. CDC’s weekly COVID-19 surveillance summary using data collected by 85 state and local public health laboratories. These reports include age-specific seroprevalence but no details regarding sample selection, test characteristics, or confidence intervals and hence could not be used in our metaregression. |
| Utsunomiya, Japan[118] | This study tested a random sample of 742 participants and found 3 confirmed positive results among 463 adults ages 18 to 65 years; the test-adjusted prevalence for that age group was 0·65% (CI: 0·13–1·8%). No positive results were obtained for the sample of 181 adults ages 65+ years. |
| Virginia, USA[119] | Virginia Department of Health collected specimens from a random sample of 3113 participants ages 16+ during early June and estimated prevalence of 2·4%. No confidence intervals or age-specific results had been released as of September 17. |
| Vo, Italy[120] | Vo’ is a municipality of 3,300 people, nearly all of whom (87%) participated in an infection survey in late February. However, there were only 54 infections among people ages 50+, so assessing age-specific IFRs is not feasible. |
| Washoe County, Nevada, USA[121] | This study collected samples from 234 individuals on June 9-10 and obtained 5 positive IgG results. No age-specific results were reported. |
| Winston-Salem, North Carolina, USA[122] | This ongoing study has been collecting specimens from a representative sample of area residents since mid-April, and raw prevalence was characterized as *“about 10%.”* On July 28 the researchers reported that the test was not sufficiently sensitive and that a new test would be deployed henceforth. |
| Zurich, Switzerland[123] | This study analyzed specimens from 578 individuals, including 90 with prior confirmed COVID-19 infections, 177 with positive patient contacts, and 311 who were randomly selected residents of Zurich. Seroprevalence in the randomly-selected group was estimated at 3·9% using the optimized test method. |

#### H.2: Studies excluded due to accelerating outbreak

| | | Cumulative fatalities in thousands | | |
|---|---|---|---|---|
| Location | Date | Study midpoint | 4 weeks later | Change (%) |
| Los Angeles, California, USA[124] | April 10-11 | 0·265 | 1·468 | 454 |
| New York City, New York, USA[10] | March 23-April 1 | 1·066 | 14·261 | 1238 |

#### H.3 Studies excluded due to non-representative samples

##### *H.3.1 Active recruitment of participants*

| Location | Description |
|---|---|
| Luxembourg[125] | Of the 35 participants who tested positive, 19 had previously interacted with a person who was known to be infected or had a prior test for SARS-CoV-2. |
| Boise, Idaho[126] | This study was promoted during a "Crush the Curve" publicity campaign and required participants to sign up for a test. |
| Santa Clara, California, USA[127] | Participants were recruited via social media and needed to drive to the testing site. Stanford Medicine subsequently released a statement indicating that the study was under review due to concerns about potential biases.[128] |
| Frankfurt, Germany[129] | This study was conducted at an industrial worksite. Among the 5 seropositive participants, 3 had prior positive tests or direct contact with a known positive case. |

##### *H.3.2 Studies of hospitals, urgent care clinics, and dialysis centers*

| Location | Description |
|---|---|
| Brooklyn, New York, USA[130] | This study used samples from an outpatient clinic and yielded a much higher infection rate than other seroprevalence studies of the New York metropolitan area. |
| Kobe, Japan[131] | This study tested for IgG antibodies in 1,000 specimens from an outpatient clinic and found 33 positive cases. However, the study did not screen out samples from patients who were seeking treatment for COVID-related symptoms. Moreover, the study reported raw prevalence and confidence interval but did not report statistics adjusted for test characteristics. The manufacturer (ADS Biotec / Kurabo Japan) has indicated that this test has specificity of 100%, based on a sample of 14 pre-COVID specimens, but that specificity has not been evaluated by any independent study. The authors concluded by noting the selection bias and recommended that *"further serological studies targeting randomly selected people in Kobe City could clarify this potential limitation."* |
| Tokyo, Japan[132, 133] | The authors of this study specifically cautioned against interpreting their results as representative of the general population. In particular, the sample of 1,071 participants included 175 healthcare workers, 332 individuals who had experienced a fever in the past four months, 45 individuals who had previously taken a PCR test, and 9 people living with a COVID-positive cohabitant. The study obtained a raw infection rate of 3·8%, but the rate is only 0·8% if those subgroups are excluded. |
| United States[134] | This study analyzed residual plasma of 28503 randomly selected adult patients receiving dialysis in July 2020. Seroprevalence for the tested sample was 8·0% (CI: 7·7–8·4%), However, prevalence of dialysis patients can diverge markedly from that of the general population; for example, this sudy finds a seropositive rate of 34% for patients residing in New York state, nearly three times higher than the 14% seroprevalence rate obtained using a random sample of that state.[11] |
| Zurich, Switzerland[135] | This study analyzed two distinct set of samples: (i) blood donors and (ii) hospital patients. Nearly all blood donors were ages 20 to 55, so that sample is not useful for assessing age-specific IFRs for older adults. The sample of hospital patients was not screened to eliminate cases directly related to COVID-19, so that sample may not be representative of the broader population, e.g., inhabitants of the city of Zurich constituted a relatively large fraction of seropositive results compared to residents from the rest of the canton of Zurich. The study found an overall IFR of 0·5% similar to that of Geneva. |

##### *H.3.3 Studies of blood donors*

| Location | Description |
| --- | --- |
| Apulia, Italy[136] | This study assessed specimens from a sample of 904 healthy blood donors at a transfusion center in southeastern Italy and obtained 9 positive results (0·99%). |
| Canada[137] | Canadian Blood Services analyzed 37737 specimens from blood donors collected between 9 May and 18 June 2020 and found 275 positive results. Test-adjusted seroprevalence was 0·7% (CI:0·60–0·79%). |
| Denmark[138] | This study assessed specimens from a sample of 20640 Danish blood donors and calculated a test-adjusted prevalence of 1·9% (CI:0·8–2·3). Unfortunately, the antibody test used in this study was subsequently identified as unreliable, and the Danish government returned all remaining test kits to the manufacturer.[139] |
| England[140] | Public Health England has conducted ongoing surveillance of seroprevalence using specimens from healthy adult blood donors. For example, in 7694 samples tested during May (weeks 18-21), the test-adjusted prevalence was 8·5% (CI: 6·9–10%). |
| Germany[141] | This study assessed residue sera from 3186 regular blood donors collected during March 9–June 3 and obtained 29 positive results (raw prevalence 0·9%). The authors stated: *"It should be emphasized that the preselection of blood donors as a study cohort is accompanied by limitations regarding representation of population."* |
| Lombardy, Italy[142] | This study assessed specimens from 390 blood donors residing in the Lodi red zone collected on April 6 and found a raw seroprevalence rate of 23%. |
| Milan, Italy[143] | This study assessed specimens from a random sample of 789 blood donors over the period from February 24 (at the start of the outbreak) to April 8. |
| Netherlands[144] | This study assessed specimens from 7361 adult blood donors collected on April 1-15 and found seroprevalence of 2·7%. |
| Rhode Island, USA[145] | This study assessed specimens from 2008 blood donors collected during April 27–May 11 and found seroprevalence of 0·6%. |
| Scotland[146] | This study assessed specimens from 3500 blood donors collected between March 17 and May 19. The authors noted that the resulting estimates of seroprevalence *"are complicated by non-uniform sampling...based on the locations where weekly donations took place...[and] further confounded by the absence of samples from individuals below age 18 and individuals over age 75."* |
| San Francisco, California, USA[147] | This study assessed specimens from 1000 blood donors that were collected during March and found one positive result (raw prevalence 0·1%). |
| United States[148] | This study analyzed residual sera from 252882 U.S. blood donors obtained between June 1 and July 31 and found an overall seroprevalence of 1·83%. |
| United States[149] | This study analyzed residual sera from 953926 U.S. blood donors obtained between June 15 and August 23 and found seropositivity of 1·82% (CI: 1·79–1·84%). |

##### *H.3.4 Studies of elementary schools*

| Location | Description |
|---|---|
| Oisie, France[150] | This sample of 1,340 participants included elementary school teachers, pupils, and their families. Only two individuals in the sample were ages 65 years and above. |
| Saxony, Germany[151] | This study analyzed specimen samples from students and teachers at thirteen secondary schools in eastern Saxony and found very low seroprevalence (0·6%). |
| Southwest Germany[76] | This study analyzed specimens from 2482 children (ages 1–10 years) and one of their parents collected between 22 April and 15 May 2020. Seroprevalence for children was 0·6% (CI: 0·3–1·0%), while the prevalence among their parents was 1·8% (CI: 1·2–2·4%). |
| Zurich, Switzerland[152] | This study collected samples from 2585 school children ages 6–16 years and found seroprevalence of 2·8% (CI: 1·6–4·1%). |

##### *H.3.5 Life insurance applicants*

| Location | Description |
|---|---|
| United States[153] | This study analyzed specimens for 50130 consecutive life insurance applicants whose blood samples were collected for insurance underwriting purposes between 12 May and 25 June 2020. The study found 1520 positive results, that is, raw seroprevalence of 3·0%. The study did not find significant differences in prevalence across three age groups (18–40, 41–60, and 61–85 years). |

##### *H.3.5 Close contacts of positive cases*

| Location | Description |
|---|---|
| Lombardy, Italy[154] | This study used a database of 62881 contacts of COVID-19 cases and conducted RT-PCR tests and antibody screening on 5484 individuals. The study reported that 2824 individuals had positive tests (51·5% of the sample), of which 62 individuals subsequently died with a COVID-19 diagnosis. |

#### H.4 Exclusion of observations with seroprevalence indistinguishable from zero

*Note: The metaregression analysis excludes observations for which either (a) the lower bound of the 95% confidence interval equals zero, and hence the upper bound of the IFR is not well defined; or (b) the lower bound of the 95% confidence interval is less than the observed COVID-19 mortality rate for that age group, implying an upper bound for the IFR that exceeds 100%.*

##### *H.4.1 Exclusion of observations from European seroprevalence studies*

| | | | 95% Confidence Interval (%) | |
|---|---|---|---|---|
| **Location** | **Age Group** | **Prevalence (%)** | **Lower Bound** | **Upper Bound** |
| Hungary[17] | 14–39 | 0·1 | 0·0 | 0·5 |
| | 40–64 | 0·3 | 0·0 | 0·6 |
| | 65+ | 0·4 | 0·0 | 0·9 |
| Portugal[18] | 20–39 | 0·9 | 0·1 | 5·3 |

##### *H.4.2 Exclusion of observations from U.S. seroprevalence studies*

| | | | 95% Confidence Interval (%) | |
|---|---|---|---|---|
| **Location** | **Age Group** | **Prevalence (%)** | **Lower Bound** | **Upper Bound** |
| Atlanta, Georgia, USA[155] | 0–17 | 0 | 0 | 1·0 |
| | 65+ | 0·7 | 0·1 | 4·5 |
| Connecticut, USA[10] | 0–18 | 0·8 | 0 | 2·9 |
| Louisiana, USA[10] | 0–18 | 2·8 | 0 | 11·5 |
| Miami, Florida, USA[10] | 0–18 | 2·4 | 0 | 7·8 |
| Minneapolis, Minnesota, USA[10] | 0–18 | 5·8 | 0 | 14·3 |
| | 50–59 | 0·7 | 0 | 2·8 |
| | 60+ | 1·0 | 0 | 3·2 |
| Missouri, USA[10] | 0–18 | 1·4 | 0 | 4·1 |
| Oregon, USA[156] | 0–17 | 0 | 0 | 10·4 |
| | 18–49 | 0 | 0 | 0·7 |
| | 50–64 | 0·1 | 0 | 1·0 |
| | 65+ | 1·4 | 0·1 | 2·8 |
| Philadelphia, Pennsylvania, USA[10] | 0–18 | 2·2 | 0 | 6·9 |
| | 50–64 | 0·8 | 0 | 2·8 |
| | 65+ | 1·6 | 0·3 | 3·5 |
| Salt Lake City, Utah, USA[14] | 65+ | 0·6 | 0 | 1·4 |
| San Francisco, California, USA[10] | 0–18 | 1·7 | 0 | 7·7 |
| | 19–49 | 1·1 | 0 | 2·6 |
| | 50–64 | 0·7 | 0 | 2·4 |
| Seattle, Washington, USA[10] | 0–18 | 0·7 | 0 | 2·5 |

##### H.5 Exclusion of observations with no observed fatalities

| Location | Age Group | Population *(millions)* | **Infections** *(thousands)* Estimate | 95% confidence interval |
|---|---|---|---|---|
| Geneva | 5–19 | 0·796 | 7·300 | 4·300–11·200 |
| Hungary | 0–14 | 1·391 | 7·795 | 3·758–11·971 |
| Portugal | 0–9 | 0·841 | 17·663 | 6·729–45·418 |
| | 10–19 | 1·015 | 21·318 | 8·121–55·834 |
| Australia | 0–39 | 13·533 | 2·800 | 4·200–12·600 |
| Iceland | 0–29 | 0·136 | 0·554 | 0·407–0·608 |
| Korea | 0–29 | 15·623 | 15·180 | 6·939–21·685 |
| Lithuania | 0–39 | 1·198 | 1·845 | 0·843–2·635 |
| New Zealand | 0–59 | 3·751 | 3·726 | 1·876–5·627 |

*Note: This table shows observations for relatively young age groups in locations where no COVID-19 fatalities were recorded for that age group and hence the implied value of the infection fatality rate is at its lower bound of zero and its confidence interval cannot be precisely determined.*

#### Appendix I: Seroprevalence Rates for Studies Included in Meta-Analysis

##### I.1 European seroprevalence studies with representative samples

| Location | Dates in 2020 | Age Group, years | Population, millions | Prevalence (%) | 95% Confidence Interval (%) |
|---|---|---|---|---|---|
| England[157] | April 16–July 3 | 0–17 | 12·023 | 9·2 | 6·2–12·2 |
| England[15] | June 20–July 13 | 18–24 | 4·747 | 7·9 | 7·3–8·5 |
| | | 25–34 | 7·609 | 7·8 | 7·4–8·3 |
| | | 35–44 | 7·147 | 6·1 | 5·7–6·6 |
| | | 45–54 | 7·623 | 6·4 | 6·0–6·9 |
| | | 55–64 | 6·782 | 5·9 | 5·5–6·4 |
| | | 65–74 | 5·576 | 3·2 | 2·8–3·6 |
| | | 75+ | 4·778 | 3·3 | 2·9–3·8 |
| France[25] | May 4–24 | 0–39 | 11·669 | 12·0 | 10·6–13·5 |
| | | 40–49 | 3·124 | 12·7 | 11·4–14·0 |
| | | 50–59 | 3·097 | 6·5 | 5·5–7·4 |
| | | 60–69 | 2·691 | 4·5 | 3·8–5·3 |
| | | 70+ | 3·209 | 3·4 | 2·7–4·1 |
| Ireland[26] | June 22–July 16 | 15–44 | 1·971 | 1·5 | 0·3–2·7 |
| | | 45–64 | 1·218 | 1·2 | 0·3–2·1 |
| Italy[16] | July 6–27 | 0–19 | 10·859 | 1·8 | 1·3–2·4 |
| | | 20–29 | 6·201 | 1·7 | 1·3–2·0 |
| | | 30–49 | 16·317 | 2·0 | 1·7–2·4 |
| | | 50–59 | 9·352 | 2·7 | 2·3–3·1 |
| | | 60–69 | 7·337 | 2·2 | 1·7–2·5 |
| | | 70+ | 10·278 | 2·1 | 1·7–2·5 |
| Netherlands[158] | April 1–17 | 0–49 | 10·053 | 3·5 | 2·5–5·2 |
| | | 50–59 | 2·524 | 4·3 | 3·2–5·8 |
| | | 60–69 | 2·130 | 3·5 | 2·5–5·0 |
| | | 70–79 | 1·592 | 3·0 | 1·7–5·3 |
| | | 80+ | 0·837 | 2·8 | 0·9–7·3 |
| Portugal[18] | May 21–July 8 | 0–9 | 0·841 | 2·2 | 0·9–6·0 |
| | | 10–19 | 1·015 | 2·4 | 0·9–6·1 |
| | | 20–39 | 2·289 | 0·9 | 0·1–5·3 |
| | | 40–59 | 3·057 | 2·6 | 1·0–6·6 |
| | | 60+ | 2·995 | 2·7 | 1·2–5·4 |
| Spain[41] | May 18–June 1 | 0–9 | 4·284 | 2·4 | 1·1–4·8 |
| | | 10–19 | 4·955 | 2·9 | 2·0–4·1 |
| | | 20–29 | 4·883 | 3·4 | 2·4–4·8 |
| | | 30–39 | 5·902 | 3·1 | 2·2–4·1 |
| | | 40–49 | 7·938 | 3·6 | 2·8–4·5 |
| | | 50–59 | 7·046 | 3·7 | 2·8–4·7 |
| | | 60–69 | 5·340 | 3·4 | 2·5–4·5 |
| | | 70+ | 6·939 | 3·1 | 1·9–5·0 |
| Geneva, Switzerland[39] | April 13–May 8 | 5–19 | 0·080 | 9·2 | 5·4–14·1 |
| | | 20–49 | 0·219 | 13·1 | 9·8–17·0 |
| | | 50–64 | 0·099 | 10·5 | 7·3–14·1 |
| | | 65+ | 0·084 | 6·8 | 3·8–10·5 |

#### I.2 European and Canadian seroprevalence studies with convenience samples

| Location | Dates in 2020 | Age Group *(years)* | Population *(millions)* | Prevalence (%) | 95% Confidence Interval (%) |
|---|---|---|---|---|---|
| Belgium[33] | April 20–26 | 0–24 | 3·229 | 6·7 | 4·7–9·6 |
| | | 25–44 | 2·957 | 6·6 | 4·7–9·2 |
| | | 45–64 | 3·081 | 6·9 | 5·2–9·2 |
| | | 65–74 | 1·147 | 4·6 | 2·6–8·0 |
| | | 75–84 | 0·691 | 7·8 | 4·7–13·0 |
| | | 85+ | 0·327 | 14·7 | 9·9–21·8 |
| Ontario[159] | June 5–30 | 0–19 | 3·142 | 0·8 | 0·3–1·4 |
| | | 20–59 | 7·977 | 1·0 | 0·7–1·3 |
| | | 60+ | 3·448 | 1·6 | 1·1–2·1 |
| Sweden[46] | April 27–May 24 | 0–19 | 2·321 | 5·7 | 4·5–7·0 |
| | | 20–49 | 3·861 | 6·5 | 5·2–7·8 |
| | | 50–69 | 2·390 | 4·8 | 3·6–6·0 |
| | | 70+ | 1·526 | 3·1 | 2·1–4·1 |

##### I.3 U.S. seroprevalence studies with representative samples

| Location | Dates in 2020 | Age Group *(years)* | Population *(millions)* | Prevalence (%) | 95% Confidence Interval (%) |
|---|---|---|---|---|---|
| Atlanta, USA[155] | April 28–May 3 | 0–17 | 0·402 | 0·0 | 0·0–1·0 |
| | | 18–49 | 0·867 | 3·3 | 1·6–6·4 |
| | | 50–64 | 0·328 | 4·9 | 1·8–12·9 |
| | | 65+ | 0·226 | 0·7 | 0·1–4·5 |
| Indiana, USA[19] | April 25–29 | 0–39 | 3·546 | 2·7 | 1·5–3·9 |
| | | 40–59 | 1·674 | 2·8 | 1·4–4·2 |
| | | 60+ | 1·512 | 1·3 | 0·6–2·0 |
| New York, USA[11] | April 23 | 0–19 | 4·898 | 14·6 | 13·1–16·1 |
| | | 20–39 | 5·409 | 14·6 | 13·1–16·1 |
| | | 40–49 | 2·356 | 15·3 | 13·7–17·0 |
| | | 50–59 | 2·623 | 16·0 | 14·6–17·5 |
| | | 60+ | 4·544 | 12·1 | 11·2–13·1 |
| Salt Lake City, USA[160, 161] | May 4–June 10 | 0–44 | 1·544 | 1·2 | 0·4–2·5 |
| | | 45–64 | 0·427 | 0·9 | 0·2–2·1 |
| | | 65+ | 0·223 | 0·6 | 0·0–1·4 |

#### I.4 U.S. Seroprevalence Studies with Convenience Samples

| Location | Dates in 2020 | Age Group *(years)* | Population *(millions)* | Prevalence (%) | 95% Confidence Interval (%) |
|---|---|---|---|---|---|
| Connecticut, USA[10] | April 26–May3 | 0·830 | 0–19 | 0·8 | 0·0–2·9 |
| | | 1·341 | 20–49 | 6·1 | 3·1–9·3 |
| | | 0·519 | 50–59 | 8·1 | 4·8– |
| | | 0·876 | 60+ | 4·2 | 2·3–6·0 |
| Louisiana, USA[10] | April 1–8 | 1·146 | 0–18 | 2·8 | 0·0–11·5 |
| | | 1·879 | 19–49 | 7·4 | 4·7–10·0 |
| | | 0·587 | 50–59 | 8·3 | 4·5–11·9 |
| | | 1·037 | 60+ | 4·4 | 1·5–8·0 |
| Miami, USA[10] | April 6–10 | 1·341 | 0–19 | 2·4 | 0·0–7·8 |
| | | 2·513 | 20–49 | 0·9 | 0·2–2·2 |
| | | 1·272 | 50–59 | 2·0 | 0·3–4·0 |
| | | 1·203 | 60+ | 3·0 | 1·7–4·5 |
| Minneapolis, USA[10] | April 30–May 12 | 0·966 | 0–18 | 5·8 | 0·0–14·3 |
| | | 1·610 | 19–49 | 2·3 | 0·8–4·2 |
| | | 0·513 | 50–59 | 0·7 | 0·0–2·8 |
| | | 0·809 | 60+ | 1·0 | 0·0–3·2 |
| Missouri, USA[10] | April 20–26 | 1·527 | 0–19 | 1·4 | 0·0–4·1 |
| | | 2·348 | 20–49 | 3·4 | 1·4–5·5 |
| | | 0·796 | 50–59 | 2·0 | 0·5–3·8 |
| | | 1·466 | 60+ | 3·2 | 1·9–4·6 |
| Philadelphia, USA[10] | April 13–25 | 0·944 | 0–18 | 2·2 | 0·0–6·9 |
| | | 1·707 | 19–49 | 5·9 | 2·4–9·8 |
| | | 1·268 | 50–59 | 0·8 | 0·0–2·8 |
| | | 0·825 | 60+ | 1·6 | 0·3–3·5 |
| San Francisco, USA[10] | April 23–27 | 1·649 | 0–18 | 1·7 | 0·0–7·7 |
| | | 2·960 | 19–49 | 1·1 | 0·0–2·6 |
| | | 1·262 | 50–59 | 0·7 | 0·0–2·4 |
| | | 1·023 | 60+ | 0·9 | 0·2–2·5 |
| Seattle, USA[10] | March 23–April 1 | 1·009 | 0–19 | 0·7 | 0·0–2·5 |
| | | 1·332 | 20–39 | 1·3 | 0·7–2·3 |
| | | 1·115 | 40–59 | 0·9 | 0·3–1·9 |
| | | 0·870 | 60+ | 1·7 | 0·9–2·7 |

**I.5 Prevalence in countries with comprehensive tracing programs**

| Location | Dates in 2020 | Population, millions | Age Group, years | Prevalence (%) | 95% Confidence Interval (%) |
|---|---|---|---|---|---|
| Australia[60] | February 1–June 12 | 13·533 | 0–39 | 0·06 | 0·03–0·09 |
| | | 6·414 | 40–59 | 0·06 | 0·04–0·10 |
| | | 2·651 | 60–69 | 0·09 | 0·05–0·13 |
| | | 1·846 | 70–79 | 0·08 | 0·04–0·12 |
| | | 1·055 | 80+ | 0·05 | 0·03–0·07 |
| Iceland[31] | February 1–June 15 | 0·136 | 0–29 | 0·4 | 0·3–0·5 |
| | | 0·132 | 30–59 | 1·1 | 0·8–1·6 |
| | | 0·038 | 60–69 | 0·5 | 0·3–1·0 |
| | | 0·023 | 70–79 | 0·3 | 0·27–1·3 |
| | | 0·013 | 80+ | 0·2 | 0·1–2·5 |
| Korea[61] | February 1–May 17 | 15·623 | 0–29 | 0·11 | 0·05–0·17 |
| | | 7·080 | 30–39 | 0·08 | 0·04–0·12 |
| | | 8·219 | 40–49 | 0·06 | 0·03–0·09 |
| | | 8·477 | 50–59 | 0·07 | 0·03–0·10 |
| | | 6·454 | 60–69 | 0·05 | 0·03–0·08 |
| | | 3·560 | 70–79 | 0·05 | 0·03–0·07 |
| | | 1·856 | 80+ | 0·06 | 0·03–0·09 |
| Lithuania[62] | February 1–June 18 | 1·198 | 0–39 | 0·15 | 0·07–0·22 |
| | | 0·356 | 40–49 | 0·18 | 0·10–0·29 |
| | | 0·421 | 50–59 | 0·17 | 0·10–0·33 |
| | | 0·353 | 60–69 | 0·13 | 0·08–0·20 |
| | | 0·223 | 70–79 | 0·09 | 0·05–0·14 |
| | | 0·172 | 80+ | 0·13 | 0·07–0·19 |
| New Zealand[63] | February 1–July 9 | 3·751 | 0–59 | 0·10 | 0·05–0·15 |
| | | 0·522 | 60–69 | 0·07 | 0·04–0·10 |
| | | 0·362 | 70–79 | 0·04 | 0·02–0·07 |
| | | 0·187 | 80+ | 0·04 | 0·02–0·06 |

**I.6 Prevalence estimates of large-scale studies used in out-of-sample analysis**

| Location | Dates in 2020 | Population, millions | Age Group, years | Prevalence (%) | 95% Confidence Interval (%) |
|---|---|---|---|---|---|
| England (ONS) | April 26–July 26 | 25·051 | 15–49 | 6·1 | 4·9–7·5 |
| | | 15·738 | 50–69 | 4·8 | 4.0–5·8 |
| | | 8·782 | 70+ | 3·9 | 3·0–5·2 |
| France[162] | April 6–12 | 0-9 | 7·527 | 5·9 | 1·6–10·2 |
| | | 10-19 | 7·883 | 3·5 | 0·7–6·4 |
| | | 20-29 | 7·371 | 7·0 | 3·8–10·2 |
| | | 30-39 | 8·011 | 3·4 | 1·0–5·8 |
| | | 40-49 | 8·326 | 7·7 | 4·6–10·9 |
| | | 50-59 | 8·635 | 9·7 | 6·4–13·1 |
| | | 60-69 | 7·765 | 10·0 | 6·5–13·5 |
| | | 70–79 | 5·728 | 5·9 | 3·1–8·7 |
| | | 80+ | 4·027 | 7·3 | 4·2–10·3 |
| Great Britain (U.K. Biobank) | May 27–July 6 | 23·562 | 0–29 | 10·8 | 9·4–12·3 |
| | | 8·641 | 30–39 | 8·2 | 7·2–9·3 |
| | | 8·180 | 40–49 | 7·2 | 6·2–8·4 |
| | | 8·810 | 50–59 | 7·1 | 6·2–8·0 |
| | | 6·928 | 60–69 | 6·4 | 5·6–7·2 |
| | | 8·782 | 70+ | 5·4 | 4·7–6·1 |
| Utah, USA (CDC) | April 20–May 3 | 1·228 | 19–44 | 1·8 | 0·6–3·5 |
| | | 0·632 | 45–64 | 2·9 | 0·9–5·2 |
| | | 0·366 | 65+ | 2·7 | 0·9–5·0 |

**I.7 Prevalence estimates of small-scale studies used in out-of-sample analysis**

| Location | Dates in 2020 | Population, | Age Group, years | Prevalence (%) | 95% Confidence Interval (%) |
|---|---|---|---|---|---|
| Diamond Princess cruise ship | Feb 1–March 7 | **1150** | 0–49 | 8·3 | NA |
| | | **398** | 50–59 | 14·8 | NA |
| | | **923** | 60–69 | 19·2 | NA |
| | | **1015** | 70–79 | 23·1 | NA |
| | | **216** | 80+ | 25·0 | NA |
| Castiglione d'Adda, Italy | May 18–25 | **3052** | 15–64 | 19·1 | 14·9–23·2 |
| | | **538** | 65–74 | 31·3 | 25·4–37·3 |
| | | **401** | 75–84 | 36·6 | 28·3–44·9 |
| | | **149** | 85+ | 42·1 | 31·1–53·1 |

#### Supplementary Appendix J: Assessment of risk of bias for included studies

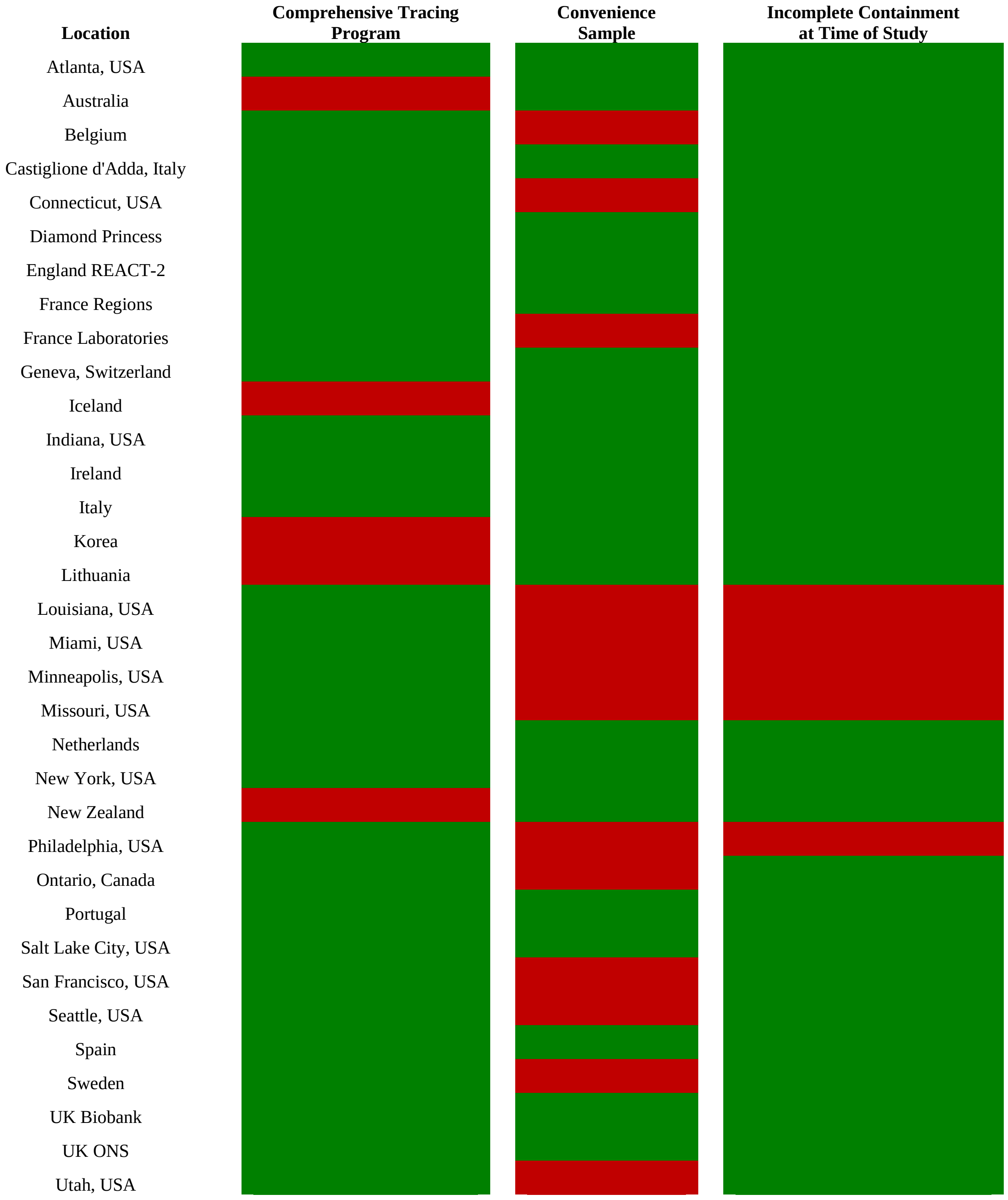

**Supplementary Appendix K: Assessment of Publication Bias**

(1) Regression-based Egger test for small-study effects
Random-effects model estimated using REML
H0: β = 0; no small-study effects
β = 0.04, SE(β) = 0.136,
z = 0.27, Prob > |z| = 0.7882

(2) Nonparametric trim-and-fill analysis of publication bias
Linear estimator, imputing on the right
Number of studies = 105
Model: Random-effects
Method: REML
Observed = 104, Imputed = 1
Effect Size and 95% Confidence Interval
Observed: 0.000 (-0.009, 0.009)
Observed + Imputed: 0.000 (-0.009, 0.009)

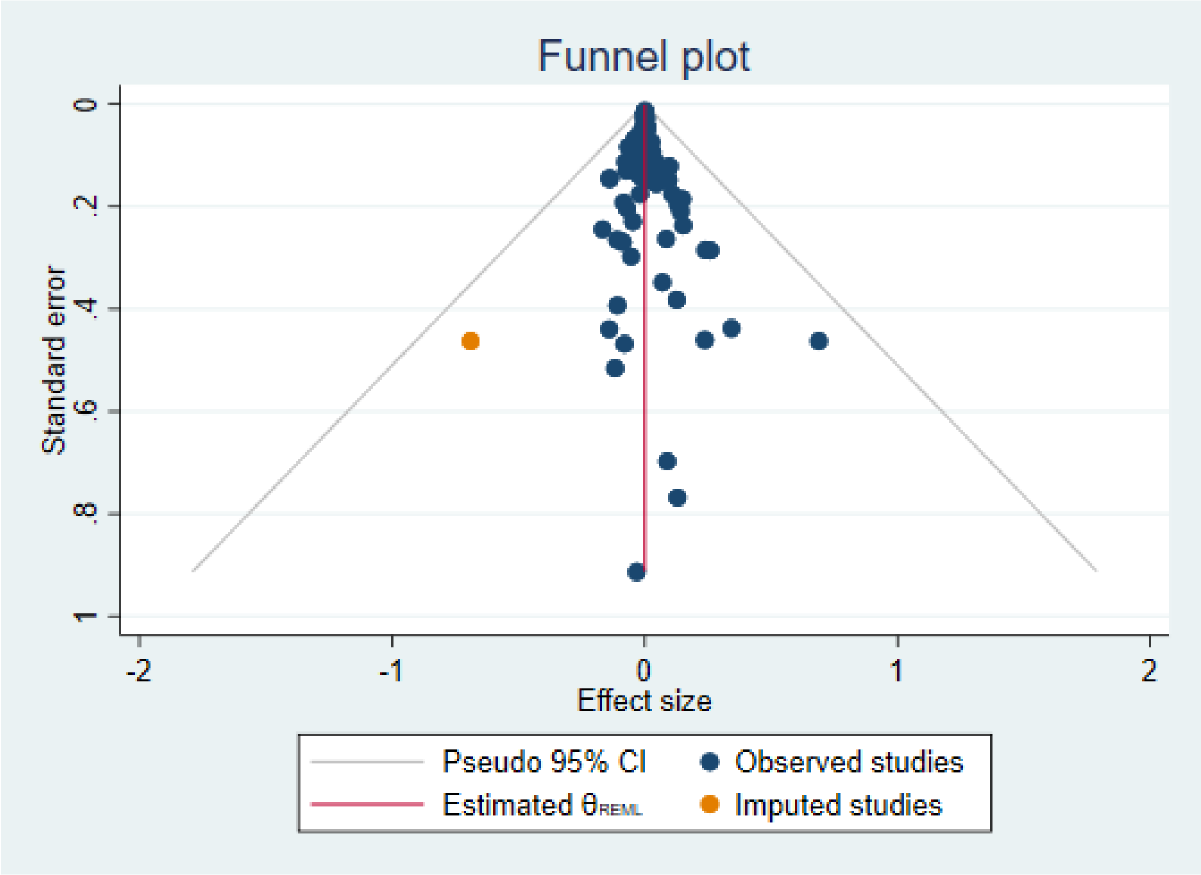

**Supplementary Appendix L: Sensitivity analysis of metaregression results**

**L.1: Stability of coefficients across age groups**

| Age Category | # Metaregression Observations | Intercept (95% CI) | Slope Coefficient (95% CI) |
|---|---|---|---|
| Age < 35 Years | 24 | -3·46 (-3·81, -3·11) | 0·065 (0·050, 0·080) |
| 35 ≤ Age ≤ 60 Years | 38 | -3·13 (-3·69, -2·57) | 0·048 (0·037, 0·060) |
| Age > 60 Years | 41 | -3·27 (-3·61, -2·93) | 0·053 (0·038, 0·067) |
| All Ages | 104 | -3·27 (-3·61, -2·93) | 0·052 (0·038, 0·067) |

**Parameter Stability Test: $H_0$ = stability of intercept and slope coefficient across age categories**
**F Statistic (4,102) = 2·13, P-value = 0·083** ***(i.e., $H_0$ is not rejected at 95% confidence level)***

**L.2: Exclusion of open-ended top age groups**

| Age Category | # Metaregression Observations | Intercept (95% CI) | Slope Coefficient (95% CI) |
|---|---|---|---|
| Benchmark Regression | 104 | -3·27 (-3·61, -2·93) | 0·053 (0·038, 0·067) |
| Exclude 60+ and 65+ | 93 | -3·26 (-3·41, -3·11) | 0·052 (0·049, 0·055) |
| Exclude 60+, 65+, 70+, and 75+ | 88 | -3·22 (-3·37, -3·07) | 0·051 (0·048, 0·054) |
| Exclude All Top Age Groups | 81 | -3·22 (-3·39, -3·06) | 0·051 (0·048, 0·054) |

#### Supplementary Appendix M: Out-of-sample analysis of metaregression results

| Study | Description |
|---|---|
| Castiglione d'Adda, Italy[163] | This study assessed seroprevalence in a random sample of 509 residents of the municipality of Castiglione d'Adda, the location of the first COVID-related fatality in Italy. Specimens were collected on May 18–25. This study is included in our meta-analysis but not in our metaregression because this municipality is covered by a nationwide seroprevalence study of Italy.[16] |
| Diamond Princess Cruise Ship[164] | This ship was carrying 3,711 passengers and crew. RT-PCR tests indicated that 619 individuals had been infected prior to the ship's debarkation on March 7, and 14 individuals subsequently died due to COVID-related causes. |
| France Laboratories[162] | Santé Publique France analyzed 3084 specimens of residual sera from two French laboratories (Cerba and Eurofins Bimnis) obtained during the week of 6–12 April, using the LuLISA-N test developed at the Institut Pasteur. Our metaregression includes a larger study of a representative sample of the French population in three regions (Grand Est, Ile de France, and Nouvelle Aquitaine) that accounted for about two-thirds of COVID-19 cases and fatalities during spring 2020. Consequently, this study is included in our meta-analysis but not in our metaregression to avoid pitfalls of nested or overlapping samples. |
| U.K. Biobank[165] | This study of Great Britain assessed seroprevalence using specimens collected from a demographically balanced panel of 17,776 participants on May 27 to July 6. Our metaregression includes a much larger seroprevalence study of the English population.[15] Consequently, this study is included in our meta-analysis but not in our metaregression to avoid pitfalls of nested or overlapping samples. |
| U.K. Office of National Statistics[13] | The U.K. Office for National Statistics (ONS) regularly reports estimates of seroprevalence from specimens provided for routine testing using an IgG ELISA test conducted by research staff at the University of Oxford. On August 18 the ONS reported age-specific results for the cumulative sample of 4840 specimens received from 26 April to 26 July and indicated that these results were broadly consistent with the findings of the UK REACT-2 study (which utilized a much larger sample). |
| Utah, USA[10] | This study analyzed commercial lab specimens from 1132 individuals collected during April 20–May 3. This study is not included in our meta-analysis because a subsequent study analyzed a much larger randomized sample of 6527 residents of the Salt Lake City metropolitan area during May 4–June 10.[160] As of May, that metro area accounted for nearly 90% of COVID-related fatalities in Utah. |

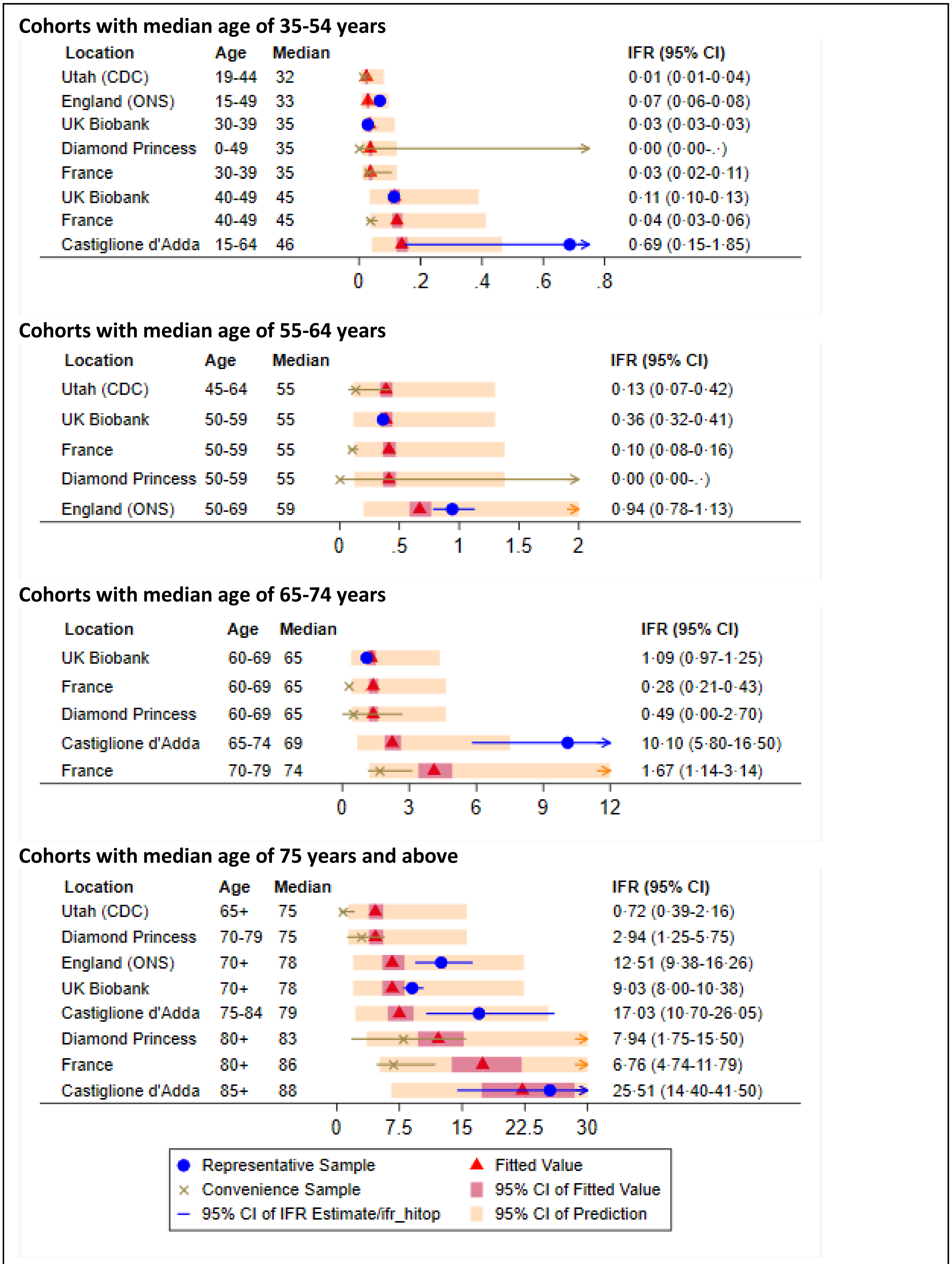

Cohorts with median age of 35-54 years
Location Age Median IFR (95% CI)
Utah (CDC) 19-44 32 0·01 (0·01-0·04)
England (ONS) 15-49 33 0·07 (0·06-0·08)
UK Biobank 30-39 35 0·03 (0·03-0·03)
Diamond Princess 0-49 35 0·00 (0·00-.·)
France 30-39 35 0·03 (0·02-0·11)
UK Biobank 40-49 45 0·11 (0·10-0·13)
France 40-49 45 0·04 (0·03-0·06)
Castiglione d'Adda 15-64 46 0·69 (0·15-1·85)
0 .2 .4 .6 .8
Cohorts with median age of 55-64 years
Location Age Median IFR (95% CI)
Utah (CDC) 45-64 55 0·13 (0·07-0·42)
UK Biobank 50-59 55 0·36 (0·32-0·41)
France 50-59 55 0·10 (0·08-0·16)
Diamond Princess 50-59 55 0·00 (0·00-.·)
England (ONS) 50-69 59 0·94 (0·78-1·13)
0 .5 1 1.5 2
Cohorts with median age of 65-74 years
Location Age Median IFR (95% CI)
UK Biobank 60-69 65 1·09 (0·97-1·25)
France 60-69 65 0·28 (0·21-0·43)
Diamond Princess 60-69 65 0·49 (0·00-2·70)
Castiglione d'Adda 65-74 69 10·10 (5·80-16·50)
France 70-79 74 1·67 (1·14-3·14)
0 3 6 9 12
Cohorts with median age of 75 years and above
Location Age Median IFR (95% CI)
Utah (CDC) 65+ 75 0·72 (0·39-2·16)
Diamond Princess 70-79 75 2·94 (1·25-5·75)
England (ONS) 70+ 78 12·51 (9·38-16·26)
UK Biobank 70+ 78 9·03 (8·00-10·38)
Castiglione d'Adda 75-84 79 17·03 (10·70-26·05)
Diamond Princess 80+ 83 7·94 (1·75-15·50)
France 80+ 86 6·76 (4·74-11·79)
Castiglione d'Adda 85+ 88 25·51 (14·40-41·50)
0 7.5 15 22.5 30
Representative Sample
Convenience Sample
95% CI of IFR Estimate/ifr_hitop
Fitted Value
95% CI of Fitted Value
95% CI of Prediction

#### Supplementary Appendix N: U.S. scenario analysis

| Scenario | Infection Rate by Age (*percent*) All | 0-49 | 50-64 | 65+ | Deaths (*thousands*) | IFR (*percent*) |
|---|---|---|---|---|---|---|
| **Scenario #1:** *uniform prevalence* | 20 | 20 | 20 | 20 | 867 | 1·3 |
| **Scenario #2:** *protection of vulnerable age groups* | 20 | 26 | 10 | 6 | 348 | 0·5 |

*Note*: These scenarios have the same average infection rate of 20% but with distinct patterns of age-specific prevalence. The metaregression estimates are used to compute total fatalities for each scenario using U.S. population data and predicted age-specific IFRs by single year of age.

#### Supplementary Appendix O: Infection fatality rate for seasonal influenza

According to the World Health Organization, seasonal influenza mortality is usually well below 0·1% while also noting that "mortality is to a large extent determined by access to and quality of health care."[166] The U.S. Center for Disease Control and Prevention provides annual estimates of the U.S. impact of seasonal influenza based on reporting of symptomatic cases from state and local public health laboratories and other sources. For the winter season of 2018-2019, its preliminary estimate is 35·5 million symptomatic cases and 34 thousand fatalities.[167] A systematic review and meta-analysis of 55 studies found that 25·4% to 61·8% of influenza infections were subclinical, i.e., did not meet the criteria for acute respiratory illness.[168] Using the midpoint of that interval, we estimate that the total U.S. incidence of seasonal influenza during winter 2018-19 was in the range of 45 million to 93 million infections and hence that the population IFR for seasonal influenza was in the range of 0.04% to 0.08%--an order of magnitude smaller than the population IFR for COVID-19.

#### Supplementary Appendix P: Excess mortality

In some locations, reported deaths may not fully capture all fatalities resulting from COVID-19 infections, especially when a large fraction of such deaths occurs outside of medical institutions. In the absence of accurate COVID-19 death counts, *excess mortality* can be computed by comparing the number of deaths for a given time period in 2020 to the average number of deaths over the comparable time period in prior calendar years, e.g., 2015 to 2019. This approach has been used to conduct systematic analysis of excess mortality in European countries.[169] For example, the Belgian study used in our metaregression computed age-specific IFRs using seroprevalence findings in conjunction with data on excess mortality in Belgium; the authors noted that Belgian excess mortality over the period from March to May coincided almost exactly with Belgium's tally of reported COVID-19 cases.[33]

**Supplementary Appendix Q: Comparison of age-specific IFRs**

| Age | Verity et al. (2020) IFR(%) | Verity et al. (2020) 95% CI | Ferguson et al. (2020) IFR (%) | Metaregression Results IFR | Metaregression Results 95% CI |
|---|---|---|---|---|---|
| 0-9 | 0·00161 | (0·0002–0·02) | 0·002 | 0·001 | (0·0007–0·0013) |
| 10-19 | 0·00695 | (0·001–0·05) | 0·006 | 0·003 | (0·002–0·004) |
| 20-29 | 0·0309 | (0·014–0·092) | 0·03 | 0·011 | (0·009–0·013) |
| 30-39 | 0·084 | (0·04–0·19) | 0·08 | 0·037 | (0·031–0·043) |
| 40-49 | 0·161 | (0·08–0·32) | 0·15 | 0·123 | (0·108–0·141) |
| 50-59 | 0·595 | (0·34–1·28) | 0·60 | 0·413 | (0·362–0·471) |
| 60-69 | 1·93 | (1·11–3·89) | 2·2 | 1·38 | (1·19–1·61) |
| 70-79 | 4·28 | (2·45–8·44) | 5·1 | 4·62 | (3·83–5·57) |
| 80+ | 7·8 | (3·80–13·3) | 9·3 | 15·46 | (12·2–19·5) |

*Note*: This table compares the metaregression results of this paper with the estimated age-specific IFRs obtained by Verity et al. (2020).[170] Those IFR estimates were subsequently adjusted by Ferguson et al. (2020) to account for non-uniform prevalence, but age-specific confidences were not reported by that study.[171]

**Supplementary Appendix R: Comparison of age-specific IFRs and CFRs**

| Age (years) | IFR (%) | CFR (%) | Ratio |
|---|---|---|---|
| 0-29 | 0·003 | 0·3 | 100 |
| 30-39 | 0·037 | 0·5 | 14·7 |
| 40-49 | 0·12 | 1·1 | 9·2 |
| 50-59 | 0·41 | 3·0 | 7·5 |
| 60-69 | 1·4 | 9·5 | 7·1 |
| 70-79 | 4·6 | 22·8 | 5·1 |
| 80+ | 15·5 | 29·6 | 2·0 |

*Note*: This table compares the estimated age-specific IFRs from our metaregression with the age-specific case fatality rates (CFRs) of Bonanad et al. (2020).[66]

**Supplementary Appendix S: Comorbidities and Demographic Factors**

While age and fatality risk are closely related, differences in the age structure of the population and age-specific infection rates surely cannot explain all deviations in IFR across regions and populations. Consequently, the role of co-morbidities and other demographic and socioeconomic factors merits further research that carefully distinguishes between infection risk and IFR.

A recent U.K. study has shown that COVID-19 mortality outcomes are strongly linked to comorbidities such as chronic pulmonary disease, diabetes, and obesity.[172] However, that study specifically warns against drawing causal conclusions from those findings, which may reflect a higher incidence of COVID-19 rather than a higher IFR for individuals with those comorbidities. Indeed, as shown in Table S1, a study of hospitalized U.K. COVID-19 patients found that patient age was far more important than any specific comorbidity in determining mortality risk.[67] For example, the COVID-19 fatality risk for an obese 40-year-old hospital patient was found to be moderately higher than for a non-obese individual of the same cohort but only one-tenth the fatality risk for a non-obese 75-year-old hospital patient.

The high prevalence of comorbidities among COVID-19 patients has been well documented but not compared systematically to the prevalence of such comorbidities in the general population. For example, one recent study of hospitalized COVID-19 patients in New York City (NYC) reported that 94% of those patients had at least one chronic health condition.[173] However, as shown in Table S2, that finding is not particularly surprising given the prevalence of comorbidities among middle-aged and elderly NYC residents. For example, nearly 30% of older NYC adults (ages 60+) are diabetic, while 23% have cardiovascular disease (including hypertension), and 8% have chronic pulmonary diseases—practically identical to the incidence of those comorbidities in the sample of hospitalized COVID-19 patients. Indeed, obesity was the *only* comorbidity that was much more prevalent among hospitalized COVID-19 patients than in the general population of older NYC adults. Nonetheless, obesity is also much more prevalent among lower-income groups who are more likely to live in high-density neighborhoods and work in high-exposure jobs, and hence such data clearly cannot be used to distinguish prevalence vs. severity of COVID-19.

Our meta-analysis has not directly considered the extent to which IFRs may vary with other demographic factors, including race and ethnicity. Fortunately, valuable insights can be garnered from other recent studies. In particular, one recent seroprevalence study of residents of two urban locations in Louisiana found no significant difference in IFRs between whites and Blacks.[99]

Nonetheless, the incidence of COVID-19 mortality among people of color is extraordinarily high due to markedly different infection rates that reflect systematic racial and ethnic disparities in housing and employment. For example, a recent infection study of a San Francisco neighborhood found that 80% of positive cases were Latinx – far higher than the proportion of Latinx residents in that neighborhood.[109] That study concluded as follows: *"Risk factors for recent infection were Latinx ethnicity, inability to shelter-in-place and maintain income, frontline service work, unemployment, and household income less than $50,000 per year."*

Other researchers have reached similar conclusions, attributing elevated infection rates among Blacks and Hispanics to dense housing of multi-generational families, increased employment in high-contact service jobs, high incidence of chronic health conditions, and lower quality of health care.[174]

In summary, while our meta-analysis has investigated the effects of age on IFR for COVID-19, further research needs to be done on how infection and fatality rates for this disease are affected by comorbidities as well as demographic and socioeconomic factors.

**Table S1: Fatality hazard ratios for hospitalized U.K. COVID-19 patients**

| Age | Hazard Ratio | Comorbidity | Hazard Ratio |
|---|---|---|---|
| 20 to 49 | 1 | Diabetes | 1·1 |
| 50 to 59 | 2·7 | Malignant Cancer | 1·1 |
| 60 to 69 | 5·5 | Chronic Cardiac Disease | 1·2 |
| 70 to 79 | 9·8 | Chronic Pulmonary Disease | 1·2 |
| 80+ | 13·5 | Chronic Kidney Disease | 1·3 |
| | | Obesity | 1·3 |
| | | Liver Disease | 1·5 |

*Source: Doherty et al. (2020), Figure 5.*

**Table S2: Comorbidity prevalence in New York City hospitalized COVID-19 patients vs. general population**

| Comorbidity | NYC Hospitalized COVID Patients | NYC Population (Ages 50+) | Difference |
|---|---|---|---|
| **Cancer** | 5·6% | 6·3% | -0·7% |
| **Cardiovascular Disease** | | | |
| Hypertension | 53·1% | 49·2% | 3·9% |
| Coronary artery disease | 10·4% | 10·5% | -0·1% |
| Congestive heart failure | 6·5% | 6·9% | -0·4% |
| **Chronic Respiratory Disease** | | | |
| Asthma | 8·4% | 8·6% | -0·2% |
| Chronic obstructive pulmonary disease | 5·0% | 7·7% | -2·7% |
| Obstructive sleep apnea | 2·7% | 2·8% | -0·1% |
| **Immunosuppression** | | | |
| HIV | 0·8% | 2·7% | -2·0% |
| History of solid organ transplant | 1·0% | NA | NA |
| **Kidney Disease** | | | |
| Chronic | 4·7% | 13·1% | -8·4% |
| End-Stage | 3·3% | 0·6% | 2·6% |
| **Liver Disease** | | | |
| Cirrhosis | 0·3% | 0·9% | -0·6% |
| Hepatitis B | 0·1% | 0·5% | -0·3% |
| Hepatitis C | 0·1% | 0·1% | 0·0% |
| **Metabolic Disease** | | | |
| Obesity (BMI>=30) | 41·7% | 26·9% | 14·8% |
| Diabetes | 31·7% | 27·6% | 4·1% |
| **Ever Smoked** | 15·6% | 43·8% | -28·2% |

*Note*: The following sources were used to gauge the prevalence of comorbidities among NYC residents ages 50 years and above. *Asthma*: U.S. Center for Disease Control & Prevention (2018). *Cancer*: New York State Cancer Registry (2016). *Cardiovascular Diseases*: New York Department of Health (2020). *Diabetes*: New York State Comptroller (2015). *HIV*: New York City Department of Health (2018). *Kidney Disease*: IPRO End-Stage Renal Disease Network of New York (2014). *Liver Disease*: Moon et al. (2019) and Must et al. (1999). *Chronic Pulmonary Disease*: New York Department of Health (2019). *Obesity*: New York City Department of Health (2019).

**Supplementary Appendix T: Metaregression Predictions for Manaus, Brazil**

A recent seroprevalence study of Manaus, Brazil found prevalence of about 66% as of August 2020; moreover, that prevalence was roughly similar across age groups.[175] As of October 29, the Brazil Ministry of Health reported 2853 confirmed COVID-19 deaths in Manaus.[176] Assuming a uniform prevalence of 66%, the following table shows the age-specific predictions of our metaregression based on the age structure of the Manaus population. The predicted death toll of 3108 is well aligned with the actual number of confirmed COVID-19 deaths, and the predicted population IFR of 0·22% is well aligned with the observed population IFR of 0·2%.

| Age Group | MedianAge | Population | Infections | Metaregression IFR (%) | Predicted Deaths |
|---|---|---|---|---|---|
| 0-9 years | 5 | 397,825 | 262,565 | 0·001 | 3 |
| 10-19 years | 15 | 431,275 | 284,642 | 0·003 | 9 |
| 20-29 years | 25 | 443,147 | 292,477 | 0·011 | 32 |
| 30-39 years | 35 | 371,428 | 245,142 | 0·037 | 90 |
| 40-49 years | 45 | 251,827 | 166,206 | 0·123 | 205 |
| 50-59 years | 55 | 156,343 | 103,186 | 0·413 | 426 |
| 60-69 years | 65 | 77,153 | 50,921 | 1·381 | 703 |
| 70+ years | 75 | 53,764 | 35,484 | 4·622 | 1,640 |
| All | 28 | 2,182,763 | 1,440,623 | 0·216 | 3,108 |

*Note:* The total population of Manaus in 2020 is reported by the Brazil Ministry of Health. The age structure of the population is assumed to be identical to that of the 2010 census, downloaded from: *https://www.citypopulation.de/en/brazil/amazonas/manaus/130260305__manaus/* .
